# Sibling Models Can Test Causal Claims without Experiments: Applications for Psychology

**DOI:** 10.1101/2025.08.25.25334395

**Authors:** S. Mason Garrison, Jonathan D. Trattner, Xuanyu Lyu, Hannah Robertson Prillaman, Laura McKinzie, Sarah H. E. Thompson, Joseph Lee Rodgers

## Abstract

Randomized experiments are the “gold standard” for inferring causation and are designed to collect covariate information. Yet, many questions cannot be answered with experiments practically or ethically. Often, potential confounds are controlled statistically as covariates in quasi-experimental designs. The typical use of covariates does not control for many systematically confounded gene-and-environmental effects. Poverty, health, and individual differences all covary with gene-and-environmental effects, so much so that using covariates can create bias. We advocate for using genetically informed designs, which strengthen causal inference by controlling for major genetic and environmental confounds even in the absence of random assignment.

We adapted the reciprocal standard dyad model into an analytic method to facilitate sibling comparisons. Differences between kin pairs explicitly distinguish within-family variance from between-family and control for all background variance linked to gene-and-environmental differences. We present four vignettes with individual differences and health outcomes. Although all four illustrations found significant associations when using covariate-based approaches, results diverged after addressing familial confounding with the discordant-kinship model. This divergence highlights the importance of considering familial influences in psychological research, demonstrating the versatility and efficacy of our method in different contexts.

---

Since Galton (1874, 1876), researchers have used twins to untangle genetic and environmental influences. Typically, such studies employ twins and other kinship data to decompose a construct’s variance into genetic and environmental variance, using behavior genetic designs (Bouchard et al., 1990; Polderman et al., 2015; Rende et al., 1990; Shields, 1958). However, those same data can be used less typically in genetically-informed quasi-experimental designs. Such designs address genetic and environmental confounds in a manner that classic methods cannot (D’Onofrio et al., 2020; Sjölander et al., 2022). Even though such confounds are pervasive in psychological research, this broad class of designs is underused in psychology (Lahey & D’Onofrio, 2010; Rowe et al., 1996; Rowe & Rodgers, 1997) can be applied to a variety of family data – beyond traditional twin models.

In this paper, we focus on kin-comparison methods for genetically-informed quasi-experimental designs. Given that twins are relatively costly to collect and may have weaknesses related to external validity, we emphasize applications with non-twin sibling data. Our three goals are to: 1) Highlight the underappreciated issues of genetic and familial confounding; 2) Advocate for broader implementation of sibling comparison designs; and 3) Illustrate the discordant-kinship model, a parsimonious regression-based method for addressing familial confounding.

## Causation in Psychology

True randomized experiments are considered ideal when inferring causation in psychology. Whenever possible, researchers ought to conduct an experiment (Cochran & Chambers, 1965; Dorn, 1953). Experiments provide a strong and straightforward basis for making causal claims under both Campbell’s (1957) and Rubin’s (1974) approaches to causal inference. Campbell’s approach to causal inference (Campbell, 1957; Shadish et al., 2002) focuses on eliminating plausible threats to validity, which is effectively addressed (on the average) with random assignment. Rubin’s counterfactual-based approach (Holland, 1988; Rubin, 1974, 2005) focuses on making verifiable assumptions. These assumptions facilitate comparing the observed outcomes under the treatment condition with the unobserved — or counterfactual — outcomes that would have occurred in the absence of the treatment. Within this framework, Rubin (2008) highlights three design features of the true experiment that contribute to its fundamental appeal: (1) treatment assignment is objective because subjects are assigned randomly to conditions, and (2) studies are "automatically designed without access to any outcome data’’ because they are inherently prospective studies;^1^ and (3) all pre-treatment covariates are “balanced” across conditions (See Diener et al., 2022; Shadish, 2010; West, 2009; West & Thoemmes, 2010 for further discussion and comparison of these approaches).

However, questions posed by psychology cannot always be answered with experiments. Often, the objective assignment to treatment is the limiting factor (i.e., not being able to randomly assign subjects to conditions for logical, ethical, or practical reasons). More broadly, if researchers are interested in:

- outcomes that are rare or unique, are long-term, or require expensive designs; or
- independent variables that cannot be manipulated, should not be manipulated, or are too expensive to be manipulated; or
- exogenous change (*i.e*., effects holding all other variables constant), rather than net effects^2^ (Todd & Wolpin, 2003),

then experiments may be impractical, unethical, or impossible to implement (West, 2009).

Instead, researchers can use quasi-experimental designs, which control for potential confounds and biases in various ways, including covariates, propensity-score matching, and instrumental variable approaches (Campbell, 1957; Rubin, 1974; Shadish et al., 2002). However, unlike randomized experiments, quasi-experimental designs (1) do not involve objectively assigned treatment;^3^ (2) are not “automatically designed” because covariates are selected based on their relationship with the outcome (Rubin, 2008); and (3) cannot guarantee that relevant pre-treatment covariates are balanced across conditions. Although all three of these distinctions are problematic, the lack of guaranteed pre-treatment group equivalence is a primary reason why many consider quasi-experimental designs are less compelling than experimental designs.

### Comparing True Experiments with the Covariate Approach

On average, random assignment handles both known and unknown biases as well as confounds. By randomly assigning participants to experimental conditions, differences within and between groups are spread out without pattern at the start of the study. Typically, the groups are equated on the average across all characteristics^4^. Thus, any differences between the groups after the experiment are logically caused by the experiment (Mill, 1846 Chapter VIII). However, random assignment cannot always be implemented; in some instances, threats to internal validity still remain after the implementation of random assignment (Shadish et al., 2002). Those include imitation of treatments, and differential mortality between the treatment and control groups. These failures occur within a study, and cannot be ruled out across replications. Another threat to internal validity is failure of randomization within a particular study. This threat occurs within a particular study as an accident of randomization (e.g., in a jumping study when by chance, the taller subjects end up in the treatment group), but is ruled out across replications. In those cases, using appropriate covariates is a standard approach to control for known biases and confounds through statistical modeling.

Using covariates is one of the most popular approaches to reducing threats to internal validity in psychology (Tracy et al., 2009) and can effectively rule out threats to internal validity in true experiments that rely on replication and “on the average” argumentation, as using covariates is effective within a given study. The covariate approach involves using variables to account for pre-treatment differences that occur absent randomization, or accidents of unfortunate randomization outcomes. When all relevant covariates are included, the covariate approach *should* eliminate those pre-treatment differences and *should* give equivalent estimates to randomized experiments across replication (Cook et al., 2009; Heckman, 1979; Mauro, 1990). In practice, however, evidence for estimate equivalence is mixed (e.g., Glazerman et al., 2003; Heckman et al., 1997; Shadish et al., 2008). Although controlling for demographic variables reduces selection bias, it often remains at considerable (and problematic) levels (Shadish et al., 2008; Steiner et al., 2010). As Lawes et al. (2025) summarize in their review of causal inference strategies, reliance on covariate adjustment—particularly in observational designs—can fail when important sources of confounding remain unmeasured or unmeasurable.

#### Understanding Familial Variance and Its Components

Regardless of the covariates used, some processes cannot be logically controlled, such as systematically confounded genetic and environmental effects (e.g., familial, Quinn et al., 2020; Rowe & Rodgers, 1997). Familial effects consist of all systematically varying genetic and environmental influences – both causal and confounding. Familial variance refers to the differences observed between families on a given trait (phenotype, dependent variable); these observable differences are influenced by genetic factors, shared-environmental factors, and potential gene-by-environment interaction effects. Collectively known as between-family variance or family-level effects, these factors encompass the common experiences and influences that, in turn, contribute to the similarities among relatives within a family.

Although behavior geneticists (like all scholars) often use shorthand definitions for "genetic" and “environmental”, these terms and their interpretation may be understood differently by general audiences (Hart et al., 2021; Visscher et al., 2008). “Genetic” effects are not literally "caused by genes;" instead, they are descriptive. Genetic variance (or heritability) reflects the proportion of variation in a trait attributable to genetic differences among individuals within a given population (Jacquard, 1983; Steinsaltz et al., 2020). Likewise, “shared-environmental” is not simply equivalent to a parenting effect—parenting can contribute to shared environmental variance but is just one of many potential influences (Hart et al., 2021; Maccoby, 2000). This variance reflects environmental influences that lead family members to be more similar to one another than genetic similarity alone would predict. These shared experiences are not limited to sharing the same home; they can also encompass the broader, shared social experience, beyond their immediate family, including shared experiences in schools, the neighborhood, friendship networks, etc. (Hart et al., 2021; Plomin & Daniels, 1987).

Yet, even these definitions include simplifications (see Bell, 1977 for a history). Technically, heritability is the “slope of the linear regression of children’s phenotypes on the mean parental phenotypes” (Jacquard, 1983; Lush, 1940). By assuming a linear relationship between phenotypic similarity and genotypic similarity, we can untangle genetic similarity from environmental sources of similarity (Falconer, 1960; Neale & Maes, 2004; Plomin et al., 1990). However, that assumption introduces complications. For instance, environments that are structured to correlate with both phenotypic and genetic similarity could also manifest as "genetic" signals (either within or across traits). A notable example is the effect of institutionalized racism on educational outcomes, where systemic barriers to educational resources lead to disparities in academic achievement. In such a society (and population), these disparities could erroneously be interpreted as evidence of genetic differences in intelligence or learning ability, when they are actually a reflection of unequal access to educational opportunities and other resources (Dar-Nimrod & Heine, 2011; Turkheimer, 2011). Regardless of the modeling assumptions used, this between-family source of variation complicates interpretation and obscures the true nature of the relationship.

### Systematic Bias

This prevalent form of systematic bias can pose challenges to interpreting associations between variables, as well as implications for interventions. Both genetic and shared-environmental confounds can obscure the true nature of relationships between variables, leading to biased estimates and potential misinterpretations. When present, familial effects bias the covariate approach in at least two ways:

1. When predictor and outcome share familial effects, familial confounds remain and artificially inflate the estimated treatment effect, creating an apparent difference between treatment and control groups that exceeds the true causal effect.
2. When covariate and outcome share familial effects, familial causes are over-controlled and are excluded from the treatment effect.

Thus, the covariate approach is particularly problematic when familial effects are present. To better understand these challenges, we can use directed acyclic graphs (DAGs), which have become increasingly valuable tools for causal inference in psychology and other social sciences (Pearl et al., 2016; Rohrer, 2018). DAGs are formal graphical representations that visually map causal assumptions and relationships between variables, with arrows denoting the direction of causality. Unlike correlation matrices or path diagrams, DAGs explicitly encode assumptions about causality and can help researchers identify both necessary controls and problematic adjustments that could induce bias.

As shown in Figure 1, DAGs can help clarify the role of familial confounding in psychological research. The key advantage of DAGs is that they force researchers to explicitly articulate their causal assumptions and identify potential sources of confounding that might not be immediately obvious in traditional statistical approaches. DAGs also help researchers avoid common pitfalls such as controlling for colliders (variables that are common effects of both the predictor and outcome; see Lawes et al. 2025, for an excellent example DAG), which can actually introduce bias rather than reduce it (Westfall & Yarkoni, 2016). To illustrate these concepts, we present two patterns of familial confounding below involving maternal depression and child outcomes.

**Figure 1.**
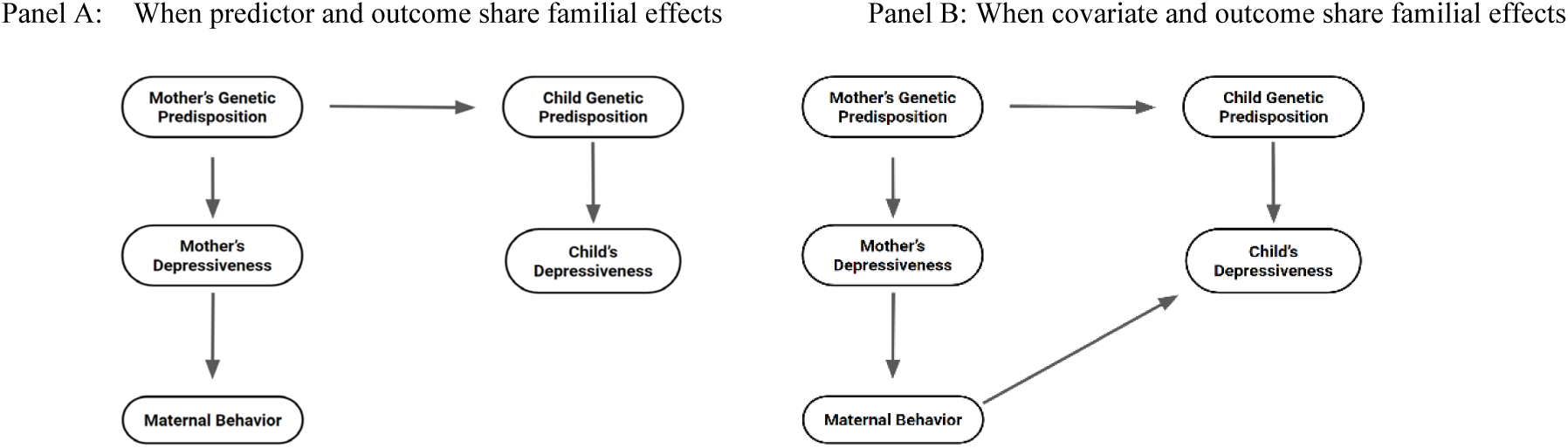
Directed Acyclic Graphs Illustrating Familial Influences in Psychological Research. *Adapted from Rohrer (2018)*. Panel A illustrates shared familial effects between predictor and outcome, where genetic predisposition represents a common cause of both maternal behavior and child depression, creating a non-causal association between them. Panel B introduces a direct causal effect of maternal behavior on child depression.

#### When predictor and outcome share familial effects (Figure 1A)

To illustrate this concept, consider the relationship between maternal parenting behavior and children’s depression. As shown in Figure 1A, this relationship is not causal but stems from familial confounding. The DAG illustrates that mothers genetically-predisposed to depression may exhibit behaviors that were historically mischaracterized as ‘refrigerator mother’ traits— perceived as cold and unloving. These genetic predispositions toward depression are inherited by children, rather than the result of any direct maternal influence. Without accounting for these shared genetic influences, researchers might incorrectly conclude that parenting style directly causes children’s depression.

#### When covariate and outcome share familial effects (Figure 1B)

Figure 1B illustrates another scenario involving maternal parenting behavior and children’s depression. Here the causal chain begins with the mother’s genetic predisposition for depression. In this scenario, mothers who have experienced depression may be more proactive in seeking treatment for their children who show signs of depression. This proactive behavior is part of the causal chain linking maternal depression and childhood depression outcomes. However, using maternal depression as a covariate in a study examining the effect of maternal behavior on childhood depression would result in over-controlling, and perhaps a misleading conclusion that maternal behavior has little to no impact on childhood depression. In reality, that maternal vigilance could substantially improve outcomes and inform targeted interventions.

DAGs help researchers distinguish between these scenarios and make informed decisions about which variables to control for and which to leave uncontrolled. Without such a framework, researchers might inadvertently introduce bias through inappropriate statistical adjustments or fail to account for important confounds. When familial influences are involved, traditional covariate approaches may be insufficient, highlighting the need for genetically-informed designs such as the sibling comparison approach detailed in this paper.

### Susceptibility to familial confounding

Although all research topics are vulnerable to systematic biases and confounds, some topics are more vulnerable. When variables with substantial univariate genetic components (i.e., moderate-to-high heritabilities) are present, there is a greater risk for systematic genetic confounds. The case is similar for traits with moderate to high environmentalities. Consequently, when both variables are incorporated into a model, the covariate approach is fundamentally biased (Rowe & Rodgers, 1997). More generally, if both associated variables are at least partially heritable^5^, their correlation may be susceptible to familial confounding. Accordingly, variables with substantial univariate genetic components (i.e., moderate-to-high heritabilities) are more prone to systematic genetic confounds. Examples of such variables include:

- personality (heritability [*h*^2^] ≈ 40%; Polderman et al., 2015; Vukasović & Bratko, 2015)
- physical health (e.g., all-cause mortality *h*^2^ ≈ 20%, weight maintenance *h*^2^ ≈ 60%, Polderman et al., 2015),
- mental health (e.g., mood disorders *h*^2^ ≈ 60%, stress reactivity *h*^2^ ≈ 30%, Polderman et al., 2015), and
- SES (*h*^2^ ≈ 50%, Garrison & Rodgers, 2019; education *h*^2^ ≈ 40%, Rietveld et al., 2013; income *h*^2^ ≈ 50%; Benjamin et al., 2012; Rowe et al., 1998).

Indeed, it is worth noting that these heritabilities are about average for what is typically seen in psychology studies (*h*^2^ ≈ 50% Polderman 2015). Other topics are not immune. but doubtless, covariate-based studies examining relationships between these variables must be interpreted with caution.

Genetic confounding occurs when the observed association between two variables is influenced by shared genetic factors, rather than a direct causal relationship between the variables themselves (Rowe & Rodgers, 1997). Similarly, shared-environmental confounding arises when environmental factors, which affect individuals in the same family, contribute to the observed association between two variables. Both types of confounding can lead to biased estimates of the relationship between variables, making it challenging to disentangle the true underlying effects.

Overlapping genetic variance reflects numerous mechanisms and thus requires content-specific interpretations. For example, the genetic overlap between neuroticism and mental health could reflect specific shared genes or nearby single-nucleotide polymorphisms (de Moor et al., 2015; Hettema, An, et al., 2006; Hettema, Neale, et al., 2006; Kendler et al., 2006; Okbay et al., 2016). Those shared genes can influence both traits through a single mechanism (such as serotonin transporter bindings, Tuominen et al., 2017) or through pleiotropy, where multiple mechanisms are influenced by the same genes. Although pleiotropy is commonly described as linking distinct phenotypes (such as physical health and neuroticism, Gale et al., 2016), pleiotropy can also link phenotypes within a general domain (e.g., generalist genes) such as learning disabilities and their respective learning abilities (Kovas & Plomin, 2006; Plomin & Kovas, 2005).

Alternatively, overlapping genetic or environmental variances could indicate proximate/direct causes, such as depression increasing later neuroticism through genetic mechanisms (Kendler et al., 1993) or prolonged exposure to stress leading to increased anxiety through environmental mechanisms (Kendler et al., 2011). These links can be more distal/indirect, such as maternal neuroticism’s impact on offspring’s mental health (Ellenbogen & Hodgins, 2004) or parenting styles impacting children’s mental health (Belsky & Pluess, 2009). Additionally, third variables with genetic or environmental variability can create a confound because of its relationship with both constructs. In such situations, the genetic or environmental variance could spill through and be detectable. For example, the genetic association between SES and physical health (Garrison & Rodgers, 2019) could be attributed to highly heritable precursors, such as intelligence (Gottfredson, 2004), which simultaneously influence both SES and physical health. Similarly, the shared environmental association between SES and mental health (Garrison & Rodgers, 2019) could reflect common environmental factors, such as neighborhood quality or access to resources, that affect both socioeconomic status and mental health outcomes by shaping individuals’ experiences, coping strategies, and social support systems. Alternatively, the association may reflect a combination of those aforementioned.

## Genetically-Informed Alternatives to Using Covariates

The most effective way to address familial confounds is to embrace them. In other words, “embracing” means to use either statistical modeling or design approaches to identify, measure, and model the nature of the confounds. Although there are numerous designs that address confounding, such as propensity-score matching and regression-discontinuity (IES, 2017), in this paper, we focus on another class of designs underused in psychology. Novel biometrical design approaches, such as children-of-twins (McAdams et al., 2014), twins-raised-apart designs (Bouchard et al., 1990), or multiple kinship pair/natural family designs (Lyu et al., 2024; Rodgers et al., 2008) can be used to explicitly address genetic confounds, using family or sibling data. Indeed, much of the field of behavior genetics uses these designs, often employing twins. Those biometrical designs can do more than merely estimate the proportion of a correlation that arises from genetic sources. They can be used to control for genetic effects within classic behavior genetic designs (e.g., Eney et al., 2017) and can be employed to test causal claims without experiments (Garrison & Rodgers, 2016; Hadd & Rodgers, 2017; Pate & Hamilton, 1992; Sims et al., 2024).

Even so, these genetically-informed designs support causal inference without random assignment through two processes: matching and Mendelian randomization (Burgess et al., 2015; Gray & Wheatley, 1991; Smith & Ebrahim, 2003). During meiosis, genetic alleles are randomly assigned to siblings, thus ensuring that all sibling background characteristics are balanced at birth, including shared environmental factors. We direct readers to the following articles for treatment on this topic — Burgess et al., 2015; Smith, 2010; Smith & Ebrahim, 2004 — as well as other applications of biology to the social sciences (Finch et al., 2001; Udry, 1995; Vogler, 2001). These processes have nuances beyond the scope of this paper. Because these biometrical designs explicitly incorporate genetic and environmental elements, they address many limitations of covariate-based designs, including many threats to internal validity. Accordingly, we advocate for their use. Yet, such models are underused in psychology (Lahey & D’Onofrio, 2010; Rodgers et al., 2000).

Classic versions of these biometrical designs rely on rare events (e.g., twins, adoptive siblings, cousin pairs) or advanced methods (e.g., genetically-informed structural equation models). These classic models impose data and knowledge-based barriers, which likely deter some researchers from using them. Many designs require specific family structures or combinations of siblings. For example, children-of-twin designs require two generations of data (D’Onofrio et al., 2003; McAdams et al., 2014), whereas studies of twins raised apart require unique circumstances (Bouchard et al., 1990). Such data can be challenging to obtain for interested researchers and typically require access to twin registries (Hur et al., 2019) or large national archival datasets with intricate kinship indicators (Rodgers et al., 2016).

Beyond the challenges of obtaining data, the most sophisticated applications of genetically-informed designs involve structural equation modeling (SEM). Although 86% of personality researchers report having used SEM at least once (Tracy et al., 2009), a survey of SEM course syllabi indicated considerable variability in content covered (Stapleton & Leite, 2005), without much coverage of behavior-genetic modeling. To bridge this gap, the Behavior Genetics Association (BGA) has sponsored specialized SEM workshops that focus on behavior-genetic modeling (Maes, 2021). Moreover, most behavior genetic applications and designs do not explicitly test questions of interest to non-behavior geneticists and instead analyze overall phenotypic variance into genetic and environmental sources. Nevertheless, although less common in practice, these techniques can be extended to explicitly test non-behavior genetics questions. In this paper, we illustrate one such technique – discordant kinship modeling.

## Discordant Kinship Modeling

To reduce these barriers, we have adapted the reciprocal standard dyad model (Kenny et al., 2001, 2006) to facilitate kin comparisons. This pooled regression approach explicitly separates within-family variance and between-family variance. It uses differences between kin pairs (*Y*_*iΔ*_ and *X*_*iΔ*_), which explicitly account for within-family variance, and kin pair means (Ȳ_*i*_ and *X̄*_*i*_), which account for between-family variance. Within-family differences create a powerful control for considerable background heterogeneity associated with both genetic and environmental differences (Lahey & D’Onofrio, 2010).

First, we predict the difference in health, *Y*_*iΔ*_, for a given kin pair (*e.g.,* twins, full siblings, half-siblings, cousins); kinship pairs are indexed as *i* in the following model:

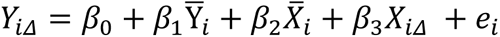

where,

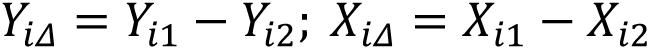

(Note that subscripts 1 and 2 identify the two individuals within the kin-pair and are defined by *Y*_*i*1_> *Y*_*i*2_. Ties are randomly assigned.)

In this model, the relative difference in kin outcome (*Y_Δ_*) is predicted from the mean level of the outcome (*Ȳ*_*i*_), the mean level of the predictor (*X̄*_*i*_), and the between-kin differences in the predictor (*X*_*iΔ*_). This approach orders siblings based on their relative outcome values (*Y*_*i*1_> *Y*_*i*2_), allowing us to extend discordant sibling analysis methods to continuous variables. Traditional discordant sibling designs typically compare siblings who differ on a binary outcome (affected/unaffected), whereas our approach extends this principle to continuous variables by treating the sibling with the higher outcome value as the "affected" sibling. This extension maintains the conceptual strength of discordant designs while broadening their application to the continuous variables commonly studied in psychological research.

This design explicitly models key explanatory processes through the explicit separation of within-family variance (with *Y*_*Δ*_ and *X*_*Δ*_) and between-family variance (with *Ȳ* and *X̄*). These mean levels, reflecting between-family variance, support causal inference by the partial controlling of genes and through the full control of the shared environment in previous generations. As long as the study is sufficiently powered, a nonsignificant association between *Y*_*Δ*_ and *X*_*Δ*_ indicates that the association between Y_1_ and X_1_ is not directly causal. Instead, it indicates that the association arose from indirect familial covariance. In contrast, a significant association between *Y*_*Δ*_ and *X*_*Δ*_ provides support for causation, with caveats.

Nonetheless, the interpretation of a significant effect depends upon the relatedness of the kin pairs involved. For instance, unlike monozygotic twins who share approximately all their genes, full siblings only share a proportion (≈ 50%) of their segregating genes. In such cases, the genetic portion of the familial covariance is attenuated, but not eliminated. This attenuation affects the power of the study to detect true causal effects. Consequently, monozygotic twin pairs provide a more powerful mechanism to support causal claims (Griliches, 1979; McGue et al., 2010; Taubman, 1976). As genetic relatedness decreases, so does the power of the design, increasing threats to internal validity.

To illustrate this point, we conducted a targeted simulation to examine the statistical power and type I error rates associated with sibling comparison analyses, specifically focused on small effect sizes (correlations ranging from .0675 to .25) and samples of 500 pairs. This parameter selection was intentional, aiming to provide a conservative estimate of power and type I error rates; in practice, many studies will have access to larger samples and may observe larger effect sizes, thus experiencing higher statistical power. This simulation, detailed in the Appendix A, found that monozygotic twins consistently showed greater power (exceeding .9) and maintained acceptable type I error rates (≈ .05), compared to full siblings. For full siblings, power and type I error rate varied as a function of how much of the association was genetic. In the absence of genetic covariance, power was lower (≈ .78), but the type I error rates were acceptable (α ≈ .05). When genetic covariance was present, power was comparable to the twin condition (>.9), but type I error rates were inflated (α ≈ .11). Nonetheless, implementing a conservative correction by requiring a significant overall F-test brought those type I error rates back to nominal levels, albeit with a slight reduction in power. Larger sample sizes will yield increased power, as well. We have provided a vignette in the discord package for users interested in conducting their own power analyses.

## Methodological Illustrations

In the current study, we highlight various aspects of the discordant-kinship design, using data from the National Longitudinal Survey of Youth 1979 (NLSY79) and the China Family Panel Survey (CFPS). Each illustration has been deliberately curated to demonstrate how different interpretations might emerge based on the analytical approach employed:

In each empirical illustration, we provide a brief theoretical motivation and then present two series of analyses. First, we present a covariate-based method and adjust for demographic variables. Then, we present those same analyses, using the discordant-kinship design. Those results are then interpreted within a brief discussion. Each illustration has a significant association when using covariate-based approaches. However, after addressing systematic familial confounding with the discordant-kinship approach, the results diverge. Although our examples apply this design to questions pertinent to differential psychology, the logic and techniques generalize broadly.

### Illustration 1 (NLSY79 Data)

Does conscientiousness causally influence physical health? This illustration demonstrates a non-causal finding by examining whether conscientiousness causally influences physical health. The discordant analysis indicates that familial factors, rather than direct causal effects, likely explain the observed association between conscientiousness and physical health.

### Illustration 2 (NLSY79 Data)

Does conscientiousness causally influence mental health? This illustration^6^ demonstrates the importance of covariates. Although the initial discordant findings suggest that conscientiousness has a causal influence on mental health, the addition of covariates shifts this interpretation from an apparent causal relationship, and no longer supports a causal conclusion.

### Illustration 3 (NLSY79 Data)

Does neuroticism causally influence mental health? This illustration demonstrates a causal effect by examining the relationship between neuroticism and mental health. This relationship remains across techniques and shows a significant effect even after applying the discordant-kinship approach with covariates.

### Illustration 4 (CFPS Data)

Does cognitive ability causally influence depression? Utilizing CFPS data, this illustration looks at the relationship between cognitive ability and depression. Its purpose is threefold: 1) to provide an unabridged results section, 2) to illustrate additional functionality present in the accompanying R package, and 3) to emphasize that these models can be applied to additional datasets.

Each illustration in this series is carefully chosen to demonstrate the varying outcomes possible when accounting for familial confounding. All analyses were conducted using R 4.5.0 (R Core Team, 2025) and the discord package (Garrison, Trattner, et al., 2025). The package includes several vignettes, including one designed to help users apply the method to their own family datasets. That vignette, which uses tools from the sister package BGmisc (Garrison et al., 2024; Garrison, Hunter, et al., 2025), demonstrates how to construct kinship links from family data, derive additive genetic and shared environmental matrices, and fit discordant-kinship models using those kinship links. More details on the kinship construction algorithm and extensive use of the Potter family pedigree can be found in Hunter, Garrison et al, 2025. Reproducible R code for each illustration is hosted on GitHub and a briefer tutorial for the accompanying provided in the supplemental materials. The NLSY illustrations are here: (github.com/R-Computing-Lab/target-causal-claims); the CFPS illustration is here (github.com/R-Computing-Lab/discord_AMPSS_CFPS).

## Methods

### NLSY Data

We analyzed data from the National Longitudinal Survey of Youth 1979 (NLSY79; described in detail in Garrison & Rodgers, 2016; Rodgers et al., 2016), which is based on a nationally-representative household probability sample. This dataset includes a broad range of psychological and health-related measures collected from 12,686 adolescents identified from 8,770 households on December 31, 1978. Respondents are surveyed biennially. Information on these publicly-available data is available in the appendix and on the Bureau of Labor Statistics (BLS) maintained website: https://nlsinfo.org/

To conduct this study using the requisite within-family information, we employed kinship pairs to support the genetically informed analysis. Our research team has completed a multi-year project to reliably and validly identify the NLSY kinship pairs (Rodgers et al., 2016) using both indirect and direct ascertainment of relatedness, resulting in a kinship classification (e.g., twins, full siblings, half-siblings, cousins, etc.) for approximately 95% (N=5,302 kinship pairs, 4006 of those full siblings) of the NLSY79 kinship pairs.

### NLSY Measures

Participants were assessed with the Ten-Item Personality Inventory (TIPI; Gosling et al., 2003), two components of SES, and two health measures. The SES components, assessed at age 50, were highest grade completed and CPI-adjusted total net family income (TNFI, Crawford et al., 2016). The two health measures, assessed at age 50, were a norm-referenced physical component summary score (PCS) from the 12-Item Short-Form Health Survey (SF-12; Ware et al., 1995), and a 7-item version of the Center for Epidemiological Studies Depression Scale (CES-D; see Levine, 2013 for psychometric properties). The complete methodology, including the reliability and application of these measures, is detailed in Appendix B. Summary statistics of these measures can be found in Table 1.

**Table 1:**
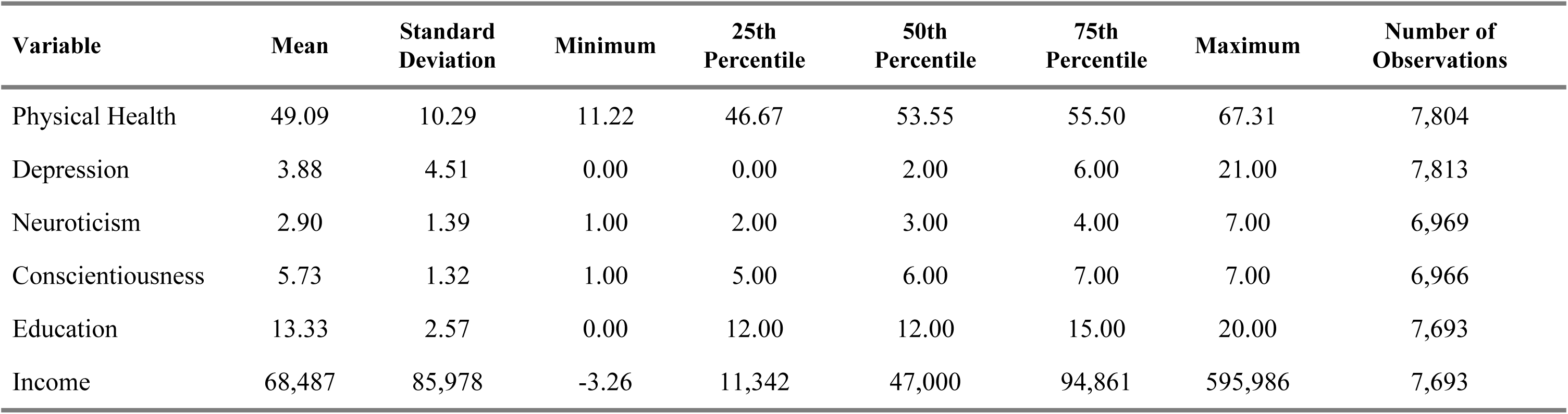
Summary Statistics for NLSY Variables as measured at age 50. Physical Health scale ranges from 10 to 70. Depression measured from CES-D, ranging from 0 to 24; Neuroticism and Conscientiousness derived from TIPI, ranging from 1 to 7; Education represents highest grade completed, ranging from 0 (none) to 20 (eight years of college or more); Income represents total net family income in 2014 dollars.

### CFPS Data

We analyzed data from the China Family Panel Survey (CFPS; Xie & Hu, 2014), a nationally-representative biennial survey of mainland Chinese households collected since 2010. As of 2018, the dataset included 58,504 individuals and extensive measures on education, health, and social relationships. The CFPS is publicly available via the Institute of Social Science Survey (ISSS) at Peking University (https://www.isss.pku.edu.cn/cfps/en/), with additional details provided in Appendix B.

To conduct within-family analyses, we identified kinship pairs using indirect and direct methods following Rodgers and colleagues (2016). This process yielded 116,909 unique relationships in the 2018 dataset, ranging from parent-child relationships to cousins (Lyu & Garrison, 2022).

### CFPS Measures

In 2018, participants completed a 24-item mathematics test to assess cognitive ability (Institute of Social Science Survey, 2010). Socioeconomic status (SES) was indexed by highest grade completed (0–23 years). Depression was measured using 9 items selected and translated from the CES-D (CES-D; Radloff, 1977; Xu et al., 2021). Numerous studies have validated the CES-D’s reliability and validity across diverse contexts (Zhang et al., 2012), cultures (Wang et al., 2013), and racial and ethnic groups (Kim et al., 2011). The complete methodology is detailed in Appendix B, and summary statistics are provided in Table 2.

**Table 2:** Summary Statistics for CFPS Variables. Math Performance ranges from 0 to 24. Depression measured from CES-D, ranging from 1 to 4; Education represents highest grade completed, ranging from 0 (none) to 23 (11 years of college or more)

| Variable | Mean | Standard<br>Deviation | Minimum | 25th<br>Percentile | 50th<br>Percentile | 75th<br>Percentile | Maximum | Number of<br>Observations |
| --- | --- | --- | --- | --- | --- | --- | --- | --- |
| Math Performance | 9.34 | 6.04 | 0 | 5 | 9.00 | 13.00 | 24 | 25,972 |
| Education | 7.71 | 4.78 | 0 | 6 | 9.00 | 12.00 | 23 | 33,974 |
| Depression | 3.32 | 0.50 | 1 | 3 | 3.38 | 3.75 | 4 | 32,598 |
| Birth Year | 1971.64 | 19.43 | 1919 | 1956 | 1970 | 1987 | 2009 | 28,419 |

### Variable Standardization

All individual-level measures were scaled to sample z-scores (*X̄*=0, SD=1) using the full sample from each dataset. For regression analyses, we fit two separate models: one using the variables standardized at the individual level (producing raw coefficients) and a second refitting approach to standardization (producing standardized coefficients). In this second approach, after computing the mean and difference scores, we standardized all variables within the analytic sample before estimating the models.

## Illustration 1: Does conscientiousness causally influence physical health?

### Brief Motivation

Conscientiousness has been consistently associated with health outcomes (Hampson, 2012; Hampson et al., 2007), including the ultimate measure of health – longevity (Chapman et al., 2010; Jackson et al., 2015; Roberts et al., 2007). Moreover, conscientiousness has also been linked with health-related behaviors (Bogg & Roberts, 2004; Friedman, 2000), which partially mediate their relationships with health (Lodi-Smith et al., 2010). Researchers have argued that personality causally influences health. Yet, plausible alternative explanations remain, partly due to the confounds that exist in the quasi-experimental nature of most personality research.

#### Analyses

Physical component scores from the SF-12 at age 50 were predicted from conscientiousness, sex, and race^7^. All measures were scaled to sample z-scores (*X̄*=0, SD=1) using the entire NLSY. In the covariate models, first-born full siblings were selected to avoid violating independence and to simplify presentation. Table 3 contains OLS regression results for the covariate-based approach, controlling for sex and race; parameter estimates are presented with standardized and raw βs with 95% confidence intervals. In the overall model, all of the independent variables explained a modest proportion of the variance in physical health scores at age 50 (F(3, 1963) = 18.50; p < 0.001; R² = 0.03).

**Table 3.** OLS: Does Conscientiousness Causally Influence Physical Health (At Age 50)?

| | Standardized<br>$\beta$ | Raw $\beta$ | t-statistic | 95% CI<sup>1</sup> | p-value |
| --- | --- | --- | --- | --- | --- |
| (Intercept) | -0.20 | -0.13 | -3.72 | -0.20, -0.06 | <0.001 |
| Conscientiousness | 0.07 | 0.06 | 2.93 | 0.02, 0.09 | 0.003 |
| Race | 0.25 | 0.23 | 5.59 | 0.15, 0.31 | <0.001 |
| Sex | 0.18 | 0.16 | 3.95 | 0.08, 0.24 | <0.001 |

<sup>1</sup>CI = Confidence Interval
$R^2 = 0.027$ ; $F(3,1963) = 18.5$ ; $p < 0.001$ ; No. Obs. = 1,967. All reported statistics and raw coefficients are from regressions where continuous variables were mean-centered and scaled by 1 SD at the individual level, and categorical predictors were dummy coded (Black or Hispanic = 0, non-Black non-Hispanic = 1; Female = 0, Male = 1). Standardized betas are from separate regressions where predictors and outcomes were fully standardized within the analytic sample prior to re-estimation.

For physical health, after adjusting for demographic variables, a standard-deviation increase in conscientiousness was associated with a .07 standard-deviation increase in health at age 50 (β = 0.07, standardized; β = 0.06, raw; t = 2.93; p = 0.003). Non-Black non-Hispanic participants had significantly higher physical health scores compared to Black or Hispanic participants (β = 0.25, standardized; β = 0.23, raw; t = 5.59; p < 0.001). Men scored higher on physical health than women (β = 0.18, standardized; β = 0.16, raw; t = 3.95; p < 0.001).

##### Discordant Analyses

Discordant results examining the association between sibling differences in conscientiousness and physical health (see Table 4). In the overall model, conscientiousness differences, physical health means, and conscientiousness means explained a substantial proportion of the variance in physical health difference (F(6, 1960) = 271.00; p < .001; R² = 0.454). Yet, sibling differences in conscientiousness were not significantly associated with sibling differences in physical health by itself (β = 0.02, standardized; β = 0.01, raw; t = 1.26; p = .209).

**Table 4.** Discordant: Does Conscientiousness Causally Influence Physical Health (At Age 50)?

|  | Standardized<br>Beta | Raw Beta | t-statistic | 95% CI<sup>1</sup> | p-value |
| --- | --- | --- | --- | --- | --- |
| (Intercept) | 0.05 | 0.921 | 29.9 | -0.015, 0.114 | <0.001 |
| Physical Health Mean | -0.67 | -0.853 | -39.3 | -0.90, -0.81 | <0.001 |
| Conscientiousness Mean | 0.04 | 0.052 | 2.45 | 0.01, 0.09 | 0.014 |
| Conscientiousness $\Delta$ | 0.02 | 0.014 | 1.26 | -0.01, 0.04 | 0.209 |
| Race | -0.05 | -0.047 | -1.49 | -0.11, 0.02 | 0.138 |
| Sex |  |  |  |  |  |
| — Sibling 1 | 0.02 | 0.016 | 0.504 | -0.05, 0.08 | 0.615 |
| — Sibling 2 | -0.07 | -0.065 | -2.09 | -0.13, 0.00 | 0.037 |

<sup>1</sup>CI = Confidence Interval
$R^2 = 0.454$ ; $F(6,1960) = 271$ ; $p < 0.001$ ; No. Obs. = 1,967. All reported statistics and raw coefficients are from regressions where continuous variables were mean-centered and scaled by 1 SD at the individual level, and categorical predictors were dummy coded (Black or Hispanic = 0, non-Black non-Hispanic = 1; Female = 0, Male = 1). Standardized betas are from separate regressions where computed predictors and outcomes were fully standardized within the analytic sample prior to re-estimation.

The coefficients of sibling-pair averages and their significance can be interpreted, but their primary purpose is to adjust for confounding. For example, sibling-pair average levels of conscientiousness were significant predictors (β = 0.04, standardized; β = 0.05, raw; t = 2.45; p = .014). A standard-deviation average increase in sibling averages in conscientiousness predicted a 0.04 standard-deviation increase in sibling differences in physical health. In other words, when siblings had higher average levels of conscientiousness, their differences in physical health were greater. Demographic variables were generally not significant, with one exception. Sibling differences in health were smaller when the less healthy sibling was male (β = −0.07, standardized; β = −0.07, raw; t = −2.09; p = .037).

#### Implications

These two analyses indicate contradictory conclusions. The covariate-based results found significant associations between conscientiousness and physical health. Thus, if presented only with covariate results, we might have concluded that this significant association suggested that conscientious behaviors lead to better health outcomes, and the relationship might well be causal. However, the discordant kinship results virtually obviate this conclusion. After accounting for familial effects, the association between conscientiousness and physical health vanished. Sibling differences in conscientiousness were not significantly associated with differences in physical health at age 50. In other words, the higher-scoring member of a sibling pair in conscientiousness was not healthier; if the link were directly causal, then we would expect the more conscientious sibling of the pair to also be healthier. Thus, the association between physical health and conscientiousness was likely not directly causal, but rather reflected familial (i.e., between-family) influences caused by external (between-family) confounds. Given that past research has shown that conscientiousness lacks shared-environmental variance (Takahashi et al., 2021; Vukasović & Bratko, 2015), we can further narrow the scope of sources of this association to genetic influences. This genetic influence could be the result of shared genes or could indicate a third variable with genetic variance. As in many findings in the social/behavioral science literature, these results suggest that past causal attributions were more likely the result of selection biases than of direct influence.

## Illustration 2 Does conscientiousness causally influence mental health?

### Brief Motivation

The association between conscientiousness and depression has been understudied relative to other Big Five traits (Klein et al., 2011). Yet, people diagnosed with major depression have reported much lower levels of conscientiousness (Cohen’s D =-1.01, Kotov et al., 2010). Moreover, conscientiousness has predicted both depressive symptoms (Chien et al., 2007) and later diagnosis of major depression (Naragon-Gainey & Watson, 2014). Because this area has been understudied, the underlying causes have not been fully specified. Some behavior genetic research has found a genetic source for the relationship between conscientiousness and lifetime diagnosis of major depression (Kendler & Myers, 2010). Whether that genetic source extends to depressive symptoms and explains the association remains an open question.

#### Analyses

Depression (using the CES-D) at age 50 was predicted using NLSY79 data from conscientiousness, highest grade completed by age 50, inflation-adjusted total net family income at age 50, sex, and race. In the covariate models, first-born full-siblings were selected to avoid violating independence and to simplify presentation. Tables 5 and 6 contain the covariate-based regression results, with and without controlling for SES component measures; parameter estimates are presented as both standardized and raw βs with 95% confidence intervals.

**Table 5.**
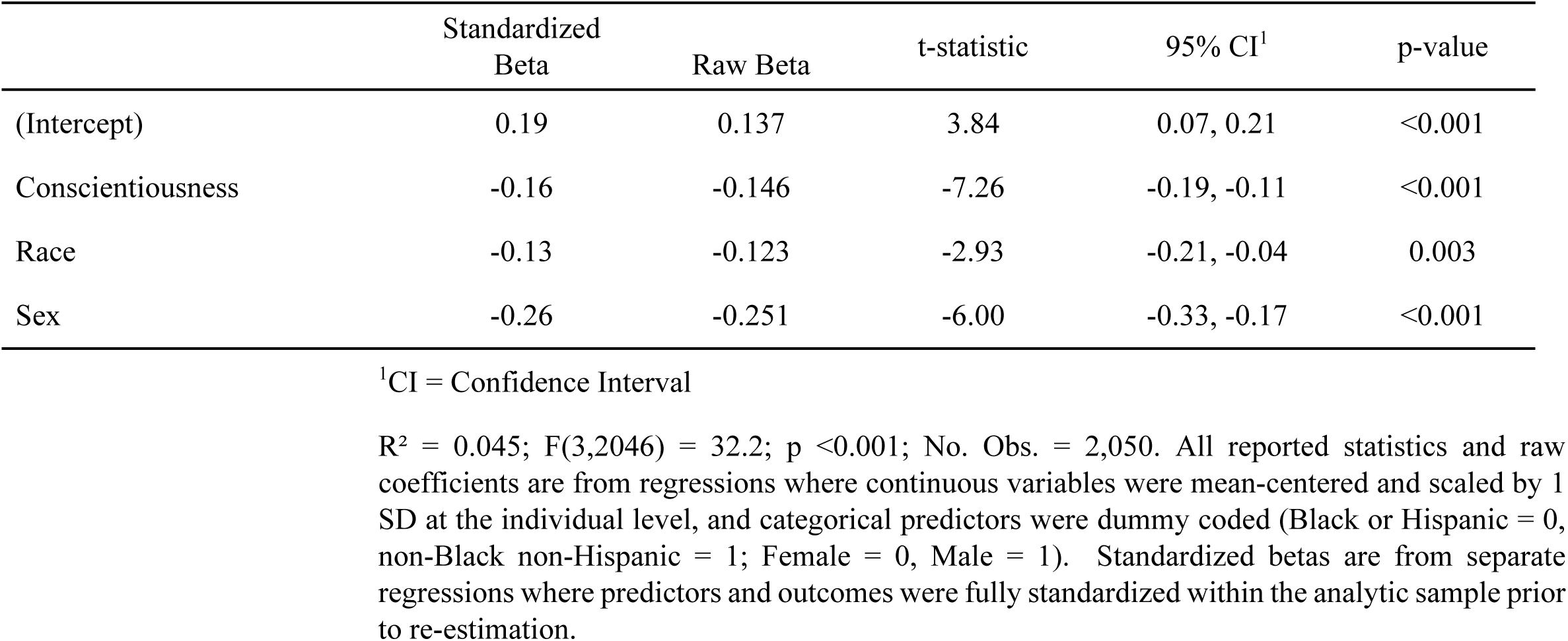
OLS: Does Conscientiousness Causally Influence Mental Health (Depression)?

|  | Standardized<br>Beta | Raw Beta | t-statistic | 95% CI<sup>1</sup> | p-value |
| --- | --- | --- | --- | --- | --- |
| (Intercept) | 0.19 | 0.137 | 3.84 | 0.07, 0.21 | <0.001 |
| Conscientiousness | -0.16 | -0.146 | -7.26 | -0.19, -0.11 | <0.001 |
| Race | -0.13 | -0.123 | -2.93 | -0.21, -0.04 | 0.003 |
| Sex | -0.26 | -0.251 | -6.00 | -0.33, -0.17 | <0.001 |

<sup>1</sup>CI = Confidence Interval
$R^2 = 0.045$ ; $F(3,2046) = 32.2$ ; $p < 0.001$ ; No. Obs. = 2,050. All reported statistics and raw coefficients are from regressions where continuous variables were mean-centered and scaled by 1 SD at the individual level, and categorical predictors were dummy coded (Black or Hispanic = 0, non-Black non-Hispanic = 1; Female = 0, Male = 1). Standardized betas are from separate regressions where predictors and outcomes were fully standardized within the analytic sample prior to re-estimation.

**Table 6.** OLS: Does Conscientiousness Causally Influence Mental Health (Depression) with Income and Highest Grade Level as Covariates?

|  | Standardized<br>Beta | Raw<br>Beta | t-statistic | 95% CI<sup>1</sup> | p-value |
| --- | --- | --- | --- | --- | --- |
| (Intercept) | 0.149 | 0.41 | 2.53 | 0.09, 0.73 | 0.011 |
| Conscientiousness | -0.138 | -0.13 | -4.72 | -0.18, -0.07 | <0.001 |
| Highest Grade at Age 50 | -0.059 | -0.056 | -1.79 | -0.118, 0.005 | 0.073 |
| TNFI at Age 50 | -0.116 | -0.100 | -3.45 | -0.156, -0.043 | <0.001 |
| Race | -0.101 | -0.09 | -1.65 | -0.20, 0.02 | 0.098 |
| Sex | -0.191 | -0.17 | -3.24 | -0.28, -0.07 | 0.001 |

<sup>1</sup>CI = Confidence Interval
$R^2 = 0.068$ ; $F(5,1117) = 16.4$ ; $p < 0.001$ ; No. Obs. = 1,123. All reported statistics and raw coefficients are from regressions where continuous variables were mean-centered and scaled by 1 SD at the individual level, and categorical predictors were dummy coded (Black or Hispanic = 0, non-Black non-Hispanic = 1; Female = 0, Male = 1). Standardized betas are from separate regressions where predictors and outcomes were fully standardized within the analytic sample prior to re-estimation.

##### Covariate Analyses

Results were consistent regardless of covariate adjustments. A standard-deviation increase in conscientiousness was significantly associated with a 0.16 standard-deviation decrease in CES-D (β = −0.16, standardized; β = −0.15, raw; t = −7.26; p < .001) without controls for SES, and a 0.14 standard-deviation decrease with controls for SES (β = −0.14, standardized; β = −0.13, raw; t = −4.72; p < .001). Men had significantly lower CES-D scores compared to women, indicating lower levels of depression (β = −0.26, standardized; β = - 0.25, raw; t = −6.00; p < .001 without SES controls, and β = −0.19, standardized; β = −0.17, raw; t = −3.24; p = 0.001 with SES controls). For highest grade completed, the results indicated a non-significant association with CES-D scores (β = −0.06, standardized; β = −0.02, raw; t = −1.79; p = 0.073). However, income was significantly associated with reduced CES-D scores in the model (β = −0.12, standardized; β = −0.10, raw; t = −3.45; p < .001). The addition of SES components increased the proportion of variance explained from 5% to 7% (R² = 0.045 without SES controls; R² = 0.068 with SES controls).

##### Discordant Analyses

Discordant results diverged, depending on covariate selection (see Tables 7 and 8). Without adjusting for SES components, sibling differences in conscientiousness were significantly associated with sibling differences in depression at age 50, controlling for conscientiousness and depression means (β = −0.04, standardized; β = −0.02, raw; t = −2.16; p = 0.031). A standard-deviation increase in the difference between siblings’ conscientiousness was associated with a 0.04 standard deviation decrease in CES-D sibling differences (note that this effect size is small, and the significant result is obtained in the context of a relatively large sample size). The negative coefficient indicated that the more conscientious sibling systematically reported lower levels of CES-D relative to their sibling. Siblings from non-Black non-Hispanic households reported a smaller difference on average than siblings from Black or Hispanic households (β = −0.10, standardized; β = −0.09, raw; t = −3.12; p = 0.002).

**Table 7.** Discordant: Does Conscientiousness Causally Influence Mental Health (Depression)?

|  | Standardized<br>Beta | Raw<br>Beta | t-statistic | 95% CI<sup>1</sup> | p-value |
| --- | --- | --- | --- | --- | --- |
| (Intercept) | 0.03 | 1.0 | 32.5 | 0.90, 1.0 | <0.001 |
| Depression Mean | 0.68 | 0.83 | 40.1 | 0.79, 0.87 | <0.001 |
| Conscientiousness Mean | 0.03 | 0.04 | 2.05 | 0.00, 0.08 | 0.041 |
| Conscientiousness $\Delta$ | -0.04 | -0.02 | -2.16 | -0.05, 0.00 | 0.031 |
| Race | -0.10 | -0.09 | -3.12 | -0.15, -0.03 | 0.002 |
| Sex |  |  |  |  |  |
| — Sibling 1 | -0.04 | -0.04 | -1.33 | -0.10, 0.02 | 0.185 |
| — Sibling 2 | 0.07 | 0.07 | 2.23 | 0.01, 0.13 | 0.026 |

<sup>1</sup>CI = Confidence Interval
$R^2 = 0.460$ ; $F(6,2043) = 290$ ; $p < 0.001$ ; No. Obs. = 2,050. All reported statistics and raw coefficients are from regressions where continuous variables were mean-centered and scaled by 1 SD at the individual level, and categorical predictors were dummy coded (Black or Hispanic = 0, non-Black non-Hispanic = 1; Female = 0, Male = 1). Standardized betas are from separate regressions where computed predictors and outcomes were fully standardized within the analytic sample prior to re-estimation.

**Table 8.**
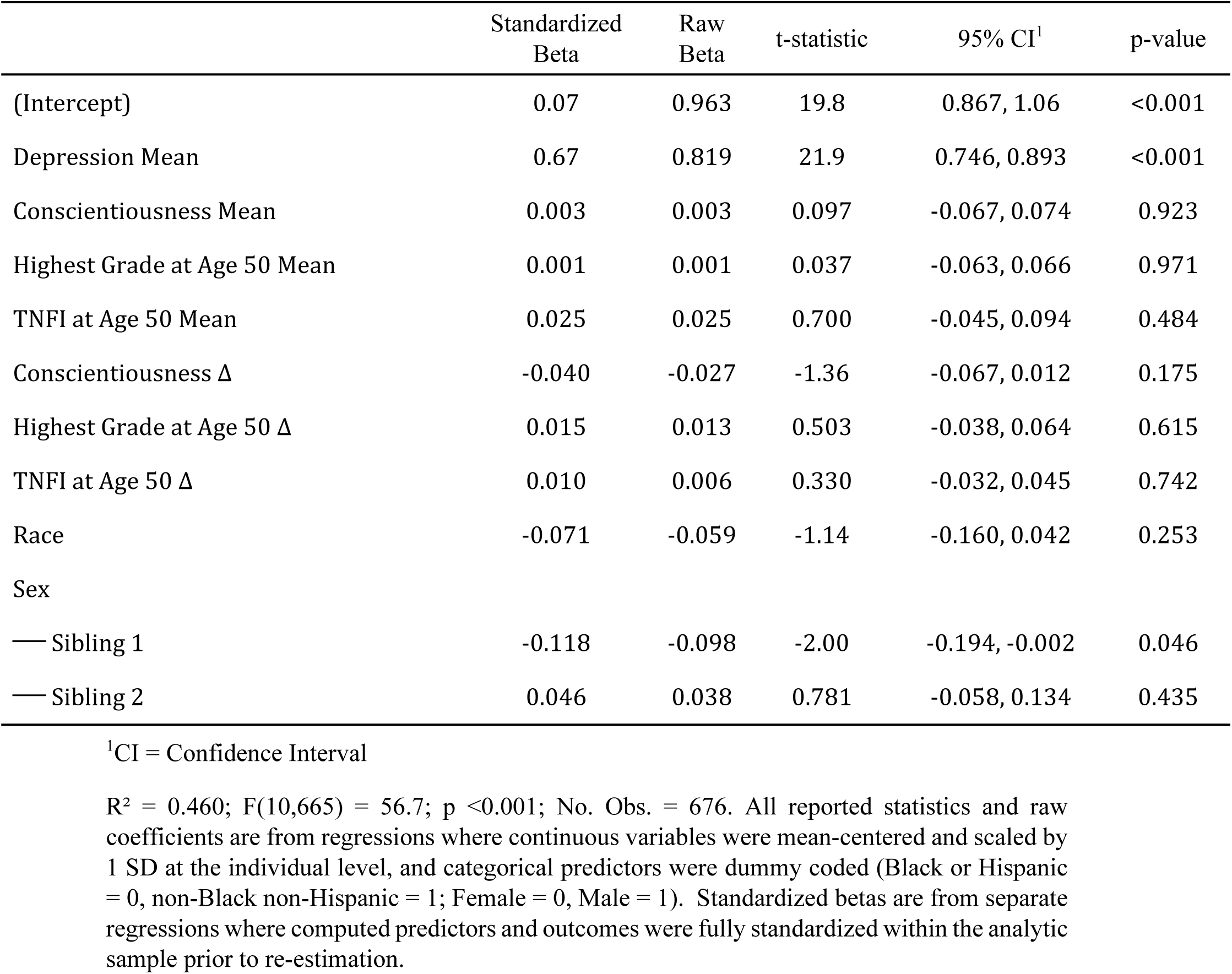
Discordant: Does Conscientiousness Causally Influence Mental Health (Depression) with Income and Highest Grade Level as Covariates?

|  | Standardized<br>Beta | Raw<br>Beta | t-statistic | 95% CI<sup>1</sup> | p-value |
| --- | --- | --- | --- | --- | --- |
| (Intercept) | 0.07 | 0.963 | 19.8 | 0.867, 1.06 | <0.001 |
| Depression Mean | 0.67 | 0.819 | 21.9 | 0.746, 0.893 | <0.001 |
| Conscientiousness Mean | 0.003 | 0.003 | 0.097 | -0.067, 0.074 | 0.923 |
| Highest Grade at Age 50 Mean | 0.001 | 0.001 | 0.037 | -0.063, 0.066 | 0.971 |
| TNFI at Age 50 Mean | 0.025 | 0.025 | 0.700 | -0.045, 0.094 | 0.484 |
| Conscientiousness $\Delta$ | -0.040 | -0.027 | -1.36 | -0.067, 0.012 | 0.175 |
| Highest Grade at Age 50 $\Delta$ | 0.015 | 0.013 | 0.503 | -0.038, 0.064 | 0.615 |
| TNFI at Age 50 $\Delta$ | 0.010 | 0.006 | 0.330 | -0.032, 0.045 | 0.742 |
| Race | -0.071 | -0.059 | -1.14 | -0.160, 0.042 | 0.253 |
| Sex |  |  |  |  |  |
| — Sibling 1 | -0.118 | -0.098 | -2.00 | -0.194, -0.002 | 0.046 |
| — Sibling 2 | 0.046 | 0.038 | 0.781 | -0.058, 0.134 | 0.435 |

<sup>1</sup>CI = Confidence Interval
$R^2 = 0.460$ ; $F(10,665) = 56.7$ ; $p < 0.001$ ; No. Obs. = 676. All reported statistics and raw coefficients are from regressions where continuous variables were mean-centered and scaled by 1 SD at the individual level, and categorical predictors were dummy coded (Black or Hispanic = 0, non-Black non-Hispanic = 1; Female = 0, Male = 1). Standardized betas are from separate regressions where computed predictors and outcomes were fully standardized within the analytic sample prior to re-estimation.

In contrast with Illustration 1, discordant results were not consistent. Specifically, after adjusting for SES components, sibling differences in conscientiousness were no longer significantly associated with sibling differences in CES-D (β = −0.04, standardized; β = −0.03, raw; t = −1.36; p = 0.175). The proportion of variance explained remained consistent across models (R² = 0.46). Moreover, sibling differences in education (β = 0.02, standardized; β = 0.01, raw; t = 0.50; p = 0.615) and income (β = 0.01, standardized; β = 0.006, raw; t = 0.33; p = 0.742) were not associated with differences in CES-D. After the inclusion of covariates, siblings from non-Black non-Hispanic households no longer reported smaller differences in CES-D relative to siblings from Black or Hispanic households (β = −0.07, standardized; β = −0.06, raw; t = −1.14; p = 0.253).

### Implications

The initial analyses suggested a consistent conclusion before adjusting for SES components. Specifically, lower levels of conscientiousness were significantly associated with higher levels of depression for both covariate-based and discordant-based approaches. Such a finding is consistent with the brief literature in this area (Klein et al., 2011). Accordingly, we may be tempted to conclude that these results are causal because we have eliminated shared-environmental effects and attenuated genetic effects. However, these initial results did not adjust for known confounds (e.g., SES, Chapman et al., 2010; Kraft et al., 2017; Roberts et al., 2007). Thus, the significant difference score could still indicate a third variable with genetic covariance or a confound in the non-shared environment (Frisell et al., 2012).

Once SES components were added to the models, the models’ conclusions diverged. In the covariate-based model, the association between conscientiousness and depression was still significant (p<.001). In contrast, in the discordant-kinship model, the association was no longer significant (p>.05). As was the case with Illustration 1, the more conscientious sibling was not the lower-scoring CES-D sibling. Had the relationship been causal, we would have expected the more conscientious member of the sibling pair to also be healthier. Thus, the association between depression symptoms and conscientiousness is likely acting through genetic pathways, perhaps mediated through SES. Although the association could result from shared genes, the SES adjustments suggest a third variable with genetic variance.

## Illustration 3 Does neuroticism causally influence mental health?

### Brief Motivation

Since ancient Greece, personality has been linked with mental health (Maher & Maher, 1994)In particular, neuroticism’s association is consistent and large, with effect sizes in the range of 0.92 (Kotov et al., 2010; Malouff et al., 2005; Ormel et al., 2013). This association potentially arises from overlapping genetic sources, which have been estimated to account for about 30-50% of the phenotypic correlation (Hettema, Neale, et al., 2006; Okbay et al., 2016), However, personality researchers argue that the relationship between mental health and neuroticism is more than merely genetic confounding, with environmental factors and causal mechanisms also playing a role (Ellenbogen & Hodgins, 2004; Jeronimus et al., 2016; Laceulle et al., 2013; Malouff et al., 2005).

#### Analyses

Mental health (CES-D) at age 50 was predicted from neuroticism, highest grade completed, inflation-adjusted TNFI, sex, and race in the NLSY79. In the covariate models, first-born full-siblings were again selected. Table 9 contains the covariate-based regression results before, adjusting for income and education; Table 10 contains the same analyses after adjusting for those covariates; parameter estimates are presented with both standardized and raw β with 95% confidence intervals. Results were relatively consistent with and without covariates. In the covariate-adjusted model, a standard-deviation increase in neuroticism was associated with a 0.16 standard-deviation increase in CES-D at age 50 (β = 0.16, standardized; β = 0.15, raw; t = 8.86; p < .001). The overall model explained 8% of the variance (R² = .08) and was significant (F(5, 1119) = 18.80, p < .001).

**Table 9.** OLS: Does Neuroticism Causally Influence Mental Health (Depression)?

|  | Standardized<br>Beta | Raw Beta | t-statistic | 95% CI<sup>1</sup> | p-value |
| --- | --- | --- | --- | --- | --- |
| (Intercept) | 0.18 | 0.14 | 3.86 | 0.07, 0.21 | <0.001 |
| Neuroticism | 0.19 | 0.19 | 8.86 | 0.14, 0.23 | <0.001 |
| Race | -0.14 | -0.14 | -3.24 | -0.22, -0.05 | 0.001 |
| Sex | -0.23 | -0.23 | -5.41 | -0.31, -0.14 | <0.001 |

<sup>1</sup>CI = Confidence Interval
$R^2 = 0.056$ ; $F(3,2042) = 40.7$ ; $p < 0.001$ ; No. Obs. = 2,046. All reported statistics and raw coefficients are from regressions where continuous variables were mean-centered and scaled by 1 SD at the individual level, and categorical predictors were dummy coded (Black or Hispanic = 0, non-Black non-Hispanic = 1; Female = 0, Male = 1). Standardized betas are from separate regressions where predictors and outcomes were fully standardized within the analytic sample prior to re-estimation.

**Table 10.** OLS: Does Neuroticism Causally Influence Mental Health (Depression) with Income and Highest Grade Level as Covariates?

|  | Standardized<br>Beta | Beta | t-statistic | 95% CI<sup>1</sup> | p-value |
| --- | --- | --- | --- | --- | --- |
| (Intercept) | 0.15 | 0.027 | 0.586 | -0.063, 0.117 | 0.558 |
| Neuroticism | 0.16 | 0.15 | 5.63 | 0.10, 0.21 | <0.001 |
| Highest Grade at Age 50 | -0.052 | -0.050 | -1.57 | -0.112, 0.012 | 0.116 |
| TNFI at Age 50 | -0.129 | -0.112 | -3.88 | -0.168, -0.055 | <0.001 |
| Race | -0.125 | -0.11 | -2.05 | -0.22, -0.005 | 0.041 |
| Sex | -0.166 | -0.151 | -2.83 | -0.256, -0.046 | 0.005 |

<sup>1</sup>CI = Confidence Interval
$R^2 = 0.078$ ; $F(5,1119) = 18.8$ ; $p < 0.001$ ; No. Obs. = 1,125. All reported statistics and raw coefficients are from regressions where continuous variables were mean-centered and scaled by 1 SD at the individual level, and categorical predictors were dummy coded (Black or Hispanic = 0, non-Black non-Hispanic = 1; Female = 0, Male = 1). Standardized betas are from separate regressions where predictors and outcomes were fully standardized within the analytic sample prior to re-estimation.

##### Discordant Analyses

Discordant kinship analyses found significant associations between sibling differences in neuroticism and sibling differences in mental health at age 50. Adjustments for various covariates did not attenuate these observed effects (see Tables 11 and 12). Covariate-adjusted results indicated that a one standard deviation increase in the difference between siblings in neuroticism was associated with a 0.11 standard deviation increase in the difference between siblings on mental health at age 50 (β = 0.11, standardized; β = 0.07, raw; t = 3.84; p < .001). The direction of this coefficient indicates that the sibling with higher neuroticism reported significantly worse mental health outcomes within the pair.

**Table 11.** Discordant: Does Neuroticism Causally Influence Mental Health (Depression)?

|  | Standardized<br>Beta | Raw<br>Beta | t-statistic | 95% CI<sup>1</sup> | p-value |
| --- | --- | --- | --- | --- | --- |
| (Intercept) | 0.03 | 0.94 | 31.9 | 0.88, 1.0 | <0.001 |
| Depression Mean | 0.67 | 0.828 | 39.1 | 0.787, 0.870 | <0.001 |
| Neuroticism Mean | -0.02 | -0.027 | -1.29 | -0.068, 0.014 | 0.198 |
| Neuroticism $\Delta$ | 0.07 | 0.046 | 4.12 | 0.024, 0.068 | <0.001 |
| Race | -0.10 | -0.093 | -3.10 | -0.151, -0.034 | 0.002 |
| Sex |  |  |  |  |  |
| — Sibling 1 | -0.03 | -0.031 | -1.04 | -0.090, 0.028 | 0.296 |
| — Sibling 2 | 0.06 | 0.059 | 1.96 | 0.000, 0.118 | 0.050 |

<sup>1</sup>CI = Confidence Interval
$R^2 = 0.463$ ; $F(6,2039) = 293$ ; $p < 0.001$ ; No. Obs. = 2,046. All reported statistics and raw coefficients are from regressions where continuous variables were mean-centered and scaled by 1 SD at the individual level, and categorical predictors were dummy coded (Black or Hispanic = 0, non-Black non-Hispanic = 1; Female = 0, Male = 1). Standardized betas are from separate regressions where computed predictors and outcomes were fully standardized within the analytic sample prior to re-estimation.

**Table 12.** Discordant: Does Neuroticism Causally Influence Mental Health (Depression) with Income and Highest Grade Level as Covariates?

|  | Standardized Beta | Raw Beta | t-statistic | 95% CI<sup>1</sup> | p-value |
| --- | --- | --- | --- | --- | --- |
| (Intercept) | 0.07 | 0.950 | 19.7 | 0.855, 1.04 | <0.001 |
| Depression Mean | 0.66 | 0.804 | 21.7 | 0.731, 0.876 | <0.001 |
| Neuroticism Mean | -0.01 | -0.001 | -0.038 | -0.070, 0.067 | 0.970 |
| Highest Grade at Age 50 Mean | 0.009 | 0.009 | 0.262 | -0.056, 0.073 | 0.793 |
| TNFI at Age 50 Mean | 0.023 | 0.022 | 0.648 | -0.046, 0.091 | 0.517 |
| Neuroticism $\Delta$ | 0.109 | 0.073 | 3.84 | 0.036, 0.110 | <0.001 |
| Highest Grade at Age 50 $\Delta$ | 0.007 | 0.006 | 0.229 | -0.044, 0.056 | 0.819 |
| TNFI at Age 50 $\Delta$ | -0.013 | -0.009 | -0.445 | -0.046, 0.029 | 0.656 |
| Race | -0.08 | -0.067 | -1.31 | -0.167, 0.033 | 0.190 |
| Sex |  |  |  |  |  |
| — Sibling 1 | -0.128 | -0.107 | -2.20 | -0.202, -0.011 | 0.028 |
| — Sibling 2 | 0.067 | 0.056 | 1.17 | -0.038, 0.151 | 0.244 |

<sup>1</sup>CI = Confidence Interval
$R^2 = 0.472$ ; $F(10,666) = 59.5$ ; $p < 0.001$ ; No. Obs. = 677 All reported statistics and raw coefficients are from regressions where continuous variables were mean-centered and scaled by 1 SD at the individual level, and categorical predictors were dummy coded (Black or Hispanic = 0, non-Black non-Hispanic = 1; Female = 0, Male = 1). Standardized betas are from separate regressions where computed predictors and outcomes were fully standardized within the analytic sample prior to re-estimation.

Other covariates in the model showed varied associations with mental health at age 50. The average depression score of siblings (β = 0.66, standardized; β = 0.80, raw; t = 21.70; p < .001) was associated with sibling differences in mental health outcomes. However, sibling averages in educational attainment at age 50 (β = 0.01, standardized; β = 0.01, raw; t = 0.262; p = 0.793) or income at age 50 (β =-0.01, standardized; β = −0.009, raw; t = −0.445; p = 0.656) were not significantly associated with differences in mental health outcomes. Neither were the sibling differences in educational attainment (β = 0.01, standardized; β = 0.01, raw; t = 0.23; p = 0.819) and family income (β = −0.01, standardized; β = −0.01, raw; t = −0.45; p = .656).

Sex and race showed mixed results. The variable sex_1 (indicating whether sibling 1 is male: β = −0.13, standardized; β = −0.11, raw; t = −2.20; p = 0.028) was negatively associated with differences in mental health outcomes, suggesting that when the sibling with more of the outcome is male, the difference between the siblings on that outcome is smaller.^8^

#### Implications

In this third illustration, the results were not contradictory. Both the covariate- and discordant-based analyses found significant associations between neuroticism and mental health. Given that we adjusted for known confounds (e.g., SES), eliminated shared-environmental effects, and attenuated genetic effects, these findings meaningfully narrow the range of alternative explanations. Although the design does not allow for definitive causal claims— residual confounding from non-shared environmental factors or incomplete genetic control may still be present—the persistence of the effect across methods makes a causal interpretation increasingly credible. A non-causal explanation remains possible, but the balance of evidence favors causation as a plausible and increasingly credible interpretation.

## Illustration 4: Does cognitive ability casually influence depression?

### Brief Motivation

Research has consistently shown that individuals with higher cognitive abilities tend to have better mental health (Jokela, 2022). Higher cognitive abilities are linked to lower rates of psychiatric disabilities (Batty et al., 2005) and fewer self-reported depressive symptoms (Khandaker et al., 2018). Indeed, lower academic achievement, often used as a proxy for cognitive ability, has been associated with higher levels of depression (Duncan et al., 2021). The relationship between cognitive ability and mental health becomes more complex when socioeconomic status (SES) is considered, particularly in adolescents. Studies have found that family SES is related to adolescents’ mental health, with depressive symptoms often more pronounced in lower SES households (Zhou et al., 2018). While studies in adulthood suggest that mental health and SES are linked through shared environmental factors (Garrison & Rodgers, 2019), adolescent-specific dynamics remain less explored. The interplay among cognitive ability, SES, and mental health likely involves multiple pathways (Jokela, 2022; Khandaker et al., 2018), and further research is needed to untangle the causal mechanisms.

#### Analyses

We employed two covariate-based regression models to explore the relationship between depression and cognitive ability in the China Family Panel Survey (CFPS). Depression was assessed using a 9-item adaptation of the CES-D. Cognitive ability was evaluated through participants’ performance on a series of mathematical problems. Our first analysis focused on individual-level data, specifically examining firstborn siblings. Additionally, we employed a between-family analysis (as was done in Garrison & Rodgers, 2016; Hadd & Rodgers, 2017; Sims et al., 2024), where sibling averages were analyzed. These analyses controlled for the highest achieved grade and birth year.

Table 13 presents the regression results for the individual-level analysis of firstborn siblings. In contrast, Table 14 contains the covariate-based regression results for the between-family analysis using sibling averages. Both tables provide parameter estimates, including standardized and raw β coefficients and 95% confidence intervals. We only interpret the between-family results for brevity, but we highlight that the results are practically identical.

**Table 13.** OLS Individual: Does cognitive ability casually influence depression?

|  | Standardized<br>Beta | Raw Beta | t-statistic | 95% CI<sup>1</sup> | p-value |
| --- | --- | --- | --- | --- | --- |
| (Intercept) | 0.000 | -9.06 | -2.29 | -16.8, -1.31 | 0.022 |
| Math Test | 0.101 | 0.063 | 2.07 | 0.003, 0.123 | 0.038 |
| Highest Grade Completed | -0.093 | -0.066 | -1.84 | -0.136, 0.004 | 0.065 |
| Birth Year | 0.075 | 0.005 | 2.44 | 0.001, 0.009 | 0.015 |

<sup>1</sup>CI = Confidence Interval
$R^2 = 0.013$ ; $F(3, 1252) = 5.65$ ; $p < 0.001$ ; No. Obs. = 1,256 All reported statistics and raw coefficients are from regressions where continuous variables were mean-centered and scaled by 1 SD at the individual level, except for Birth Year. Standardized betas are from separate regressions where predictors and outcomes were fully standardized within the analytic sample prior to re-estimation.

**Table 14.** OLS Between Family: Between Family: Does cognitive ability casually influence depression?

|  | Standardized<br>Beta | Beta | t-statistic | 95% CI<sup>1</sup> | p-value |
| --- | --- | --- | --- | --- | --- |
| (Intercept) | 0.000 | -9.81 | -2.19 | -18.6, -1.04 | 0.028 |
| Math Test (2018) Mean | 0.180 | 0.142 | 3.60 | 0.064, 0.219 | <0.001 |
| Highest Grade Completed Mean | -0.131 | -0.118 | -2.58 | -0.209, -0.028 | 0.010 |
| Birth Year Mean | 0.068 | 0.005 | 2.23 | 0.001, 0.009 | 0.026 |

<sup>1</sup>CI = Confidence Interval
$R^2 = 0.022$ ; $F(3,1252) = 9.26$ ; $p < 0.001$ ; No. Obs. = 1,256. All reported statistics and raw coefficients are from regressions where continuous variables were mean-centered and scaled by 1 SD at the individual level, except for Birth Year. Standardized betas are from separate regressions where predictors and outcomes were fully standardized within the analytic sample prior to re-estimation.

Although the overall model was statistically significant (F(3, 1252) = 9.26, p < .001), it explained a relatively small proportion of the variance in average depression scores among sibling pairs (R² =0.022). Our results indicated that for the average sibling pair, a one standard deviation increase in the math test score average was associated with a 0.18 standard deviation increase in the average depression score (β = 0.18, standardized; β = 0.142, raw; t = 3.60, p < .001), controlling for the average highest achieved grade and age.

Furthermore, sibling averages in highest achieved grade were negatively associated with sibling averages of depression (β = −0.13, standardized; β = −0.018, raw; t = −2.58, p = .010), indicating that sibling pairs with higher educational attainment, on average, reported lower levels of depression. Birth year was positively associated with average depression scores (β = 0.07, standardized; β = 0.005, raw; t = 2.23, p = .026), indicating that younger sibling pairs tended to have higher average depression scores.

##### Discordant Analyses

The discordant model was employed to examine the relationship between sibling differences in cognitive ability and sibling differences in depression, controlling for educational attainment and siblings’ birth years. Two variations of the model were analyzed: one presented in Table 15, which excluded differences in birth year and education, and another in Table 16, which included these variables. Given the similarity in results from both models, our interpretation focuses on the simpler model presented in Table 15.

**Table 15.** Discordant: Does cognitive ability casually influence depression?

|  | Standardized<br>Beta | Raw<br>Beta | t-statistic | 95% CI<sup>1</sup> | p-value |
| --- | --- | --- | --- | --- | --- |
| (Intercept) | 0.000 | -0.991 | -0.220 | -9.82, 7.84 | 0.826 |
| Mental Health Mean | -0.370 | -0.396 | -13.9 | -0.452, -0.340 | <0.001 |
| Math Test Mean | 0.007 | 0.006 | 0.141 | -0.072, 0.084 | 0.888 |
| Highest Grade Completed Mean | -0.045 | -0.043 | -0.938 | -0.134, 0.047 | 0.349 |
| Birth Year Mean | 0.012 | 0.001 | 0.412 | -0.003, 0.005 | 0.680 |
| Math Test $\Delta$ | 0.025 | 0.016 | 0.964 | -0.017, 0.048 | 0.335 |

<sup>1</sup>CI = Confidence Interval
$R^2 = 0.138$ ; $F(5,1250) = 40.2$ ; $p < 0.001$ ; No. Obs. = 1,256. All reported statistics and raw coefficients are from regressions where continuous variables were mean-centered and scaled by 1 SD at the individual level, except for Birth Year. Standardized betas are from separate regressions where computed predictors and outcomes were fully standardized within the analytic sample prior to re-estimation.

**Table 16.** Discordant: Does cognitive ability casually influence depression (extended model)?

|  | Standardized<br>Beta | Raw<br>Beta | t-statistic | 95% CI<sup>1</sup> | p-value |
| --- | --- | --- | --- | --- | --- |
| (Intercept) | 0.00 | 0.577 | -0.129 | -9.38, 8.23 | 0.898 |
| Mental Health Mean | -0.367 | -0.392 | -13.8 | -0.447, -0.336 | <0.001 |
| Math Test Mean | 0.007 | 0.008 | 0.195 | -0.070, 0.086 | 0.846 |
| Highest Grade Completed Mean | -0.043 | -0.044 | -0.945 | -0.134, 0.047 | 0.345 |
| Birth Year Mean | 0.010 | 0.001 | 0.320 | -0.004, 0.005 | 0.749 |
| Math Test $\Delta$ | 0.072 | 0.045 | 1.78 | -0.005, 0.096 | 0.076 |
| Highest Grade Completed $\Delta$ | -0.059 | -0.045 | -1.34 | -0.110, 0.021 | 0.181 |
| Birth Year $\Delta$ | 0.052 | 0.008 | 2.20 | 0.001, 0.016 | 0.028 |

<sup>1</sup>CI = Confidence Interval
$R^2 = 0.138$ ; $F(5,1250) = 40.2$ ; $p < 0.001$ ; No. Obs. = 1,256. All reported statistics and raw coefficients are from regressions where continuous variables were mean-centered and scaled by 1 SD at the individual level, except for Birth Year. Standardized betas are from separate regressions where computed predictors and outcomes were fully standardized within the analytic sample prior to re-estimation.

The overall model explained 13.9% of the variance in sibling differences in depression scores (F(5, 1250) = 40.4, p < .001; R² = 0.139). The results indicated that differences in math test scores between siblings were not significantly associated with differences in depression scores (β = 0.025, raw; β = 0.016, t = 0.964, p = 0.335). In other words, the sibling with higher levels of depression was not also the sibling with higher level scores on math performance.

Interestingly, the average mental health status of the sibling pair emerged as a significant predictor of differences in depression scores between siblings (β = −0.370, standardized; β = - 0.396, raw, t=-13.9, p <.001). Specifically, a one-standard-deviation increase in the sibling-pair average mental health level predicted a 0.37 standard deviation decrease in the difference in depression scores between siblings. The analysis also showed that sibling averages in educational attainment (β = −0.045, standardized; β = −0.043, raw; t = −0.938; p = .349) and birth year (β = 0.012, standardized; β = 0.001, raw; t = 0.412; p = 0.680) were not significantly associated with differences in depression scores.

#### Implications

Like the first two of our other examples, these two series of analyses indicate contradictory conclusions. The covariate-based results found significant associations between mental health and cognitive ability, specifically linking lower levels of depression with higher math performance. However, the discordant kinship results virtually obviate this conclusion, showing that after accounting for familial effects, the association between mental health and cognitive ability is no longer significant. Sibling differences in depression were not significantly associated with sibling differences in math test scores. In other words, the sibling with the lower depression score did not score significantly higher on the math test than the sibling with the higher depression score; if the link were directly causal, then we would expect the sibling with better mental health to also score better in the math test. Thus, the association between mental health and cognitive ability found in the covariate-based model was likely not directly causal, but rather reflected familial (i.e., between-family) sources of variance.

#### Next Steps in Sibling Comparison Studies

The illustrations demonstrate the family of potential outcomes that researchers could encounter when using sibling comparison designs. Specifically, we demonstrated two primary outcomes: non-significant results in the sibling comparison analyses, which are strongly suggestive of non-causal relationships, and significant results in the sibling comparison analyses suggesting potential causal effects. Although each scenario presents unique challenges and opportunities, there are general strategies researchers can employ to build upon their findings.

This section briefly outlines broad recommendations for follow-up steps based on the potential outcomes of sibling comparison analyses.

For non-significant results, as demonstrated in our first, second, and fourth illustrations, the model effectively eliminates direct causal explanations. Instead it indicates that familial influences likely drive the observed relationships, and the result is not causal. Next steps to delve deeper into those familial effects likely driving the observed relationship could include:

- Attempting to distinguish between genetic and shared-environmental factors within these familial influences. For example, in our conscientiousness and physical health illustration (Illustration 1), further research could employ Cholesky modeling to parse out these components.
- Replicate the study using different kinship levels, such as adopted siblings or cousins. Our conscientiousness and mental health study (Illustration 2) could be extended using adopted siblings to further isolate environmental effects. Preregistering such a replication would clarify its confirmatory status and strengthen its interpretive value
- Improve measurement quality. The brief TIPI measure used in our NLSY79 illustrations could be replaced with more comprehensive personality inventories which would reduce the non-shared environmental variance.
- Increase sample size, especially for small effects. This is particularly relevant for our cognitive ability and depression study using CFPS data (Illustration 4), where effects may be small.
- Formalize inference around null effects using model comparison. Researchers may wish to use model selection strategies (e.g., AIC or BIC) to compare nested models, or fit constrained models in which specific coefficients are fixed to zero. These approaches can help quantify relative support for the absence of a direct effect in well-powered sibling models.

When results remain significant after controlling for familial confounds, as in our third illustration on neuroticism and mental health, researchers should consider the following steps:

- Examine the role of covariates. Carefully investigate the role of covariates in the theoretical model. The use of directed acyclic graphs (DAGs) can help clarify hypothesized causal relationships (Pearl et al., 2016). In our third illustration, for example, a DAG could help visualize the potential pathways through which neuroticism might influence mental health, accounting for the effects of SES and other relevant variables.
- Explore non-shared environmental factors. Consider potential non-shared environmental factors that might explain the persistent association. This evaluation could involve collecting more detailed data on individual-specific experiences or environments (Turkheimer & Waldron, 2000). In the case of neuroticism and mental health, this might include examining differential exposure to stressors or access to mental health resources within sibling pairs.
- Employ monozygotic twins. If possible, researchers should consider pre-registering and replicating their findings using monozygotic twins to fully eliminate genetic confounding (Vitaro et al., 2009).
- Apply advanced modeling techniques. Explore more complex models, such as multivariate Cholesky behavior genetic models, to evaluate potential mediating pathways (Loehlin, 1996).
- Pilot interventions that include all siblings. Evaluate whether effects identified in sibling comparison designs can be reproduced experimentally. Petersen and Lange (2020) mathematically proved that, when using fixed-effects OLS estimation, sibling comparison designs identify causal effects that operate at the family level, not solely at the individual level. Therefore, to test such effects— such as the causal relationship between neuroticism and mental health— intervention trials should include all siblings in a family, not just those with elevated neuroticism.

Across both paths, researchers should remember that this model is most effective at narrowing the scope of potential causes rather than confirming any single cause. The discordant kinship model’s primary strength lies in its ability to eliminate causal claims rather than affirm them. Regardless of whether results are significant or non-significant, researchers should focus on examining family effects for potential causal relationships, while addressing validity threats.

## Discussion

### Illustrations

After introducing the logic underlying genetically-informed designs and the discordant-kinship model, we presented four illustrations. In this series of illustrations, we applied both covariate-based designs and discordant-kinship designs to four distinct empirical scenarios, using individual differences and health data from the two large nationally representative datasets. All four illustrations found significant associations when using covariate-based approaches. After addressing systematic confounding with the discordant-kinship approach, illustration results diverged. The relationship between conscientiousness and physical health vanished when tested within a discordant-kinship framework and served to illustrate interpreting a nonsignificant within-family effect. The relationship between conscientiousness and depression remained until a covariate was added to the model. This illustration emphasized that a significant effect could still be the result of third variables. The third illustration found that the significant association between neuroticism and mental health remained even after adjusting for plausible confounders. The final illustration demonstrates how these models can be applied to additional datasets, examining the relationship between cognitive ability and depression in a non-Western sample.

Like our first illustration, this example found that the apparent relationship between cognitive ability and depression disappeared after accounting for familial confounding It is no accident that we used the NLSY79 to illustrate many of these principles of causal interpretation. The NLSY79 is a powerful data source for studying family-based processes, especially when combined with the genetically-informed kinship links (Rodgers et al., 2016) that allow discordant kinship and behavior genetic designs to be applied. The empirical illustrations in this paper were chosen to illustrate different outcome patterns that can occur, but other NLSY studies have shown conclusions from the research literature to be causally questionable, or in some cases, incorrect. Such studies include Garrison and Rodgers (2016) on the relationship between age at first intercourse and intelligence; Hadd & Rodgers (2017) on the relationship between maternal and child intelligence and the construction of the home environment; D’Onofrio et al. (2008) on the relationship between smoking during pregnancy and childhood problem behaviors; and Jaffee et al. (2011), on whether placing a child in daycare affects academic performance later in childhood. Rodgers et al. (2019) used different types of cousin pairs (full cousins, half cousins, quarter cousins, etc.) to identify a variance source as a potential causal influence on height outcomes that Sir Ronald Fisher had originally viewed as spurious.

Although the relationship remained, we encouraged researchers to interpret these findings as providing some degree of evidence in favor of a causal effect, given the careful consideration of other plausible confounders, a priori evidence, and theoretical plausibility. Similar to other designs, we recommend that researchers interpret these findings as suggestive, rather than conclusive, because we have not eliminated every threat to validity. Rather, the significant difference score indicates that a non-trivial association remains, after controlling for familial effects. This design is more effective at narrowing the scope of causes rather than confirming any single cause.

### What sibling designs can and cannot do

Broadly, kin comparison designs offer powerful tools for addressing many issues related to internal validity by modeling familial confounding. Added design features can further enhance kin comparison studies, but the basic design does not address other threats to statistical validity, construct validity, or external validity. Maturation is addressed if the kin are the same age (e.g., twins). History is addressed if both kin experience the event. Much of the kin comparison design’s appeal stems from its ability to partially address selection.

Selection is partially addressed and depends on the kin group as well as the source of the selection. Non-shared environmental selection is addressed by this class of designs (Frisell et al., 2012) with additional design features, such as covariates. If selection is related to the shared environment, then it is fully addressed when the kin were raised in the same home (Lahey & D’Onofrio, 2010). Epigenetic effects can only be addressed when the environmental source of the gene-by-environment interaction is shared-environmental.

Other genetic-related selection effects are partially addressed through two processes: matching and Mendelian randomization. When monozygotic twins are used, then genetic-related selection effects are matched in the same way that the shared-environment is matched (McGue et al., 2010). Note, however, that monozygotic twins can be discordant in phenotypic and genotypic expression (Fraga et al., 2005; Machin, 1996). Mendelian randomization, in contrast, partially addresses familial effects in non-identical siblings (Burgess et al., 2015; Gray & Wheatley, 1991; Smith & Ebrahim, 2003). During meiosis, genetic alleles are randomly assigned, thus ensuring that all sibling background characteristics are balanced at birth on average. Mendelian randomization does not address multi-function of genes (i.e., third variables), genetic mediation, developmental compensation (Smith & Ebrahim, 2004) or confounding in the non-shared environment (Frisell et al., 2012). Even with these caveats, the family-based selection addressed is an improvement over non-genetically informed design.

Finally, one of the strongest features of sibling comparison designs is the control for ancestral family background that they provide. Full siblings share all ancestors from their parents back, for thousands of generations. Both genetic and environmental (and interaction) differences that naturally occur among unrelated individuals are fully controlled using sibling comparisons. Further, these controls operate at the level of a single study (in which siblings are compared), and not just on the average across replications.

### Appealing design features of experiments

At the outset of this paper, we highlighted the three appealing design features of experiments (Rubin, 2008), contrasting their absence in non-experiments. We argue that the discordant kinship design possesses (most of) those three features: 1) Treatment assignment is objective. Within a kin pair, assignment is based on which kin member has more of the outcome (e.g., higher health). Theoretically, you could assign the ordering based on whichever had more of the treatment, but modeling multiple predictors complicates the assignment. 2) The design is automatic “without access to outcome data” (Rubin, 2008). Although the design is automatic, treatment assignment requires outcome data. 3) Pre-treatment covariates are balanced across conditions. Within a kin pair, all background covariates are balanced at birth (see above paragraph). This last element is both a strength of this design as well as its major limitation. After birth, the genetic influences are still balanced due to Mendelian randomization, as are the shared-environmental influences. However, as the siblings age, they accumulate non-shared experiences. These non-shared experiences are not balanced. Accordingly, spurious significant results can arise from differences in non-shared environmental influences. For non-monozygotic siblings, the design attenuates but does not fully eliminate genetic confounding within a sibling pair (though past ancestral genetic confounding is fully controlled). Thus, spurious significant results can still arise.

These two limitations do restrict the effectiveness of this model for confirming causation, but not for casting doubt upon it. This design remains a powerful, yet parsimonious technique for testing causal claims without experiments. If a causal claim cannot overcome this parsimonious test, it cannot be directly causal for any causal effect that should still be seen within a family. In contrast, when a causal claim cannot be eliminated through this test, as in the sibling with more of the outcome is also the sibling with more of the predictor, that lends provisional support to causal effects. When that information is taken together with other considerations such as a priori evidence and theoretical plausibility, it may very well be causal.

In summary, we have contrasted — both conceptually and empirically — covariate-based methods with discordant kinship designs. We have shown empirically that causal assertions from covariate-based approaches do not always match those using kinship pairs. Further, we have shown conceptually the strength of using kinship information to address causal assertions. Such methods do not eliminate certain threats to causal inference as effectively as true experiments; but they *actually* improve on experiments in certain ways; and they provide substantial advantages over many other quasi-experimental methods by reducing threats to internal validity. Because family designs control for virtually all background heterogeneity, a primary source of selection bias is substantially reduced by using these types of methods, providing evidence that can contribute to our understanding of causal effects.

## Supporting information

Simulation

Tutorial

## Data Availability

Reproducible R code for each illustration is hosted on GitHub and a briefer tutorial for the accompanying provided in the supplemental materials. The NLSY illustrations are here: (github.com/R-Computing-Lab/target-causal-claims); the CFPS illustration is here (github.com/R-Computing-Lab/discord_AMPSS_CFPS).

https://github.com/R-Computing-Lab/target-causalclaims

https://github.com/R-Computing-Lab/discord_AMPSS_CFPS

https://www.isss.pku.edu.cn/cfps/en/

https://nlsinfo.org/

## Footnotes

1 When properly conducted (e.g., using pre-registration, double-blinding), neither the null nor alternative hypothesis can be systematically favored. In contrast, when the researcher has access to information about the outcome during the design process, significant results can be produced with p-hacking (Gelman & Loken, 2014; Lakens, 2015) and harking (hypothesizing after the result is known; Kerr, 1998).

2 Exogenous change examines the direct influence of one variable on the outcome, holding all variables constant. In contrast, net effect or total effect examines the influence of manipulating one variable on the outcome, including any secondary effects (Todd & Wolpin, 2003). This distinction is discussed more often in the intervention literature and is sometimes described as structure-preserving interventions and structure-altering interventions.

3 Some quasi-experimental designs do indeed use objective treatment assignment, such as regression discontinuity.

4 These characteristics include genetic elements that may systematically correlate with an experimental condition (Lahey & D’Onofrio, 2010).

5 A corollary of this “rule” is that if one of the variables lacks a genetic underpinning, then the association between the two variables cannot arise from a genetic component.

6 Because this scenario is rare, the specific illustration is intentionally and extremely p-hacked—it took over twenty attempts to produce an empirical example that fit. We commend the reviewers for identifying the odd design choices, which are a good indication of questionable research practices (Stefan & Schönbrodt, 2023). For readers interested in non-p-hacked findings, we direct readers to Garrison & Rodgers, 2019.

7 Race was determined through a combination of self-identification, interviewer report, and inference from household reports (Ward & Burich, 1978).

8 This coefficient interpretation is not particularly informative or relevant to the research question about causation.

