## Supplementary material for "Sibling Models Can Test Causal Claims without Experiments: Applications for Psychology": Simulation

**Power Simulation**

This supplement delves deeper into the simulation discussed in the main manuscript regarding the relationship between kin-pair relatedness and the interpretation of significant effects in our model. As noted in the manuscript, the interpretation of a significant effect depends upon the relatedness of the kin pairs involved. For instance, unlike monozygotic twins who share all their genes, full siblings only share a proportion (≈ 50%) of their segregating genes. In such cases, the genetic portion of the familial covariance is attenuated, but not eliminated. This attenuation affects the power of the study to detect true causal effects. Consequently, monozygotic twin pairs provide a more powerful mechanism to support causal claims (Griliches, 1979; McGue, Osler, & Christensen, 2010; Taubman, 1976). As genetic relatedness decreases, so does the power of the design, increasing threats to validity.

To quantify these assertations, we conducted a small simulation. Specifically, we varied the following conditions with 1000 replications per condition:

- Relatedness:
  - Monozygotic Twins or Full Siblings (r): 1, .5
- Between Family Covariance:
  - Genetic Covariance: 0, .25
  - Shared Environmental Covariance: 0, .25
- Within Family Covariance:
  - Non-Shared Environmental covariance: 0, .25

In each condition, we generated 500 kin pairs and created two variables, with 25% genetic variance, 25% shared-environmental variance, and 50% non-shared environmental variance, using the discordsim function in the discord package (Garrison & Ream, 2017). The magnitude of the covariance was based on typical effect sizes of psychological constructs (Polderman et al., 2015). Accordingly, the total correlation between variables ranged from 0 to .25. Power and type I error were calculated at p=.05, as the proportion of replications that found a significant difference score. The type 1 error rate was defined as finding a significant effect when the non-shared environmental covariance was 0, whereas power was defined as finding a significant effect when the non-shared environmental covariance was .25.

We present our findings in Table 1. The monozygotic twin condition had greater power (power> .9) and lower type I error rate (α ≈ .05) than the full sibling conditions. For full siblings, power and type I error rate varied as a function of genetic covariance. In the absence of genetic covariance, power was lower (≈ .78), but the type I error rates were acceptable (α ≈ .05). In the presence of genetic covariance, power was comparable to the twin condition (>.9), but type I error rates were inflated (α ≈ .11). This inflated type 1 error rate can be corrected to nominal levels (α ≈ .05) by requiring a significant overall F test. Those corrected values are also presented in Table 1. This correction is conservative; it reduced power across all full-sibling conditions by .17 and over-corrected alpha (α ≈ .02) in the conditions without genetic covariance. Accordingly, we recommend using the conservative correction for full siblings when researchers suspect genetic influences. Throughout this paper, we will use that conservative criteria. In general, these results demonstrate that this design is highly powered, even with small correlations. Larger samples and larger effects would further improve power beyond these levels, and probably normalize type I error rates as well.

Table

*Table 1 Power and Type 1 Error rate to detect non-shared environmental covariance with the discordant kinship model*

| Conditions | | | | | Results | | |
| --- | --- | --- | --- | --- | --- | --- | --- |
| Relatedness | Genetic Covariance | Shared Environmental Covariance | Nonshared Environmental Covariance | Phenotypic Correlation | Median Observed Correlation (90 CI) | Proportion of Replications with a Significant Difference Score (and a significant F-test) | |
|  |  |  |  |  |  | Power | Empirical Alpha |
| 1 | 0 | 0 | 0 | 0 | 0 (-0.06, 0.055) |  | 0.042 (0.019) |
| 1 | 0.25 | 0 | 0 | 0.0625 | 0.066 (0.006, 0.119) |  | 0.045 (0.016) |
| 1 | 0 | 0.25 | 0 | 0.0625 | 0.062 (0.007, 0.119) |  | 0.04 (0.011) |
| 1 | 0.25 | 0.25 | 0 | 0.125 | 0.126 (0.066, 0.182) |  | 0.049 (0.021) |
| 1 | 0 | 0 | 0.25 | 0.125 | 0.124 (0.068, 0.182) | 0.917 (0.808) |  |
| 1 | 0.25 | 0 | 0.25 | 0.1875 | 0.185 (0.13, 0.242) | 0.93 (0.829) |  |
| 1 | 0 | 0.25 | 0.25 | 0.1875 | 0.188 (0.133, 0.244) | 0.929 (0.817) |  |
| 1 | 0.25 | 0.25 | 0.25 | 0.25 | 0.252 (0.196, 0.304) | 0.916 (0.823) |  |
| 0.5 | 0 | 0 | 0 | 0 | 0.001 (-0.054, 0.052) |  | 0.039 (0.015) |
| 0.5 | 0.25 | 0 | 0 | 0.0625 | 0.062 (0.004, 0.119) |  | 0.115 (0.047) |
| 0.5 | 0 | 0.25 | 0 | 0.0625 | 0.063 (0.008, 0.119) |  | 0.051 (0.016) |
| 0.5 | 0.25 | 0.25 | 0 | 0.125 | 0.126 (0.069, 0.178) |  | 0.106 (0.051) |
| 0.5 | 0 | 0 | 0.25 | 0.125 | 0.127 (0.074, 0.181) | 0.773 (0.586) |  |
| 0.5 | 0.25 | 0 | 0.25 | 0.1875 | 0.185 (0.133, 0.239) | 0.936 (0.854) |  |
| 0.5 | 0 | 0.25 | 0.25 | 0.1875 | 0.186 (0.135, 0.242) | 0.786 (0.614) |  |
| 0.5 | 0.25 | 0.25 | 0.25 | 0.25 | 0.25 (0.195, 0.299) | 0.94 (0.841) |  |

Notes: For both variables, a^2^ and c^2^ were assumed to be .25, and e^2^ to be .5. Nominal alpha was set to .05, and sample size was set to 500 pairs. 1000 replications per condition.
