## Supplementary material for "Sibling Models Can Test Causal Claims without Experiments: Applications for Psychology": Data Appendix

**Data Appendix B**

### National Longitudinal Survey of Youth 1979 (NLSY79)

#### **Data**

The National Longitudinal Survey of Youth 1979 (NLSY79; described in detail in Garrison & Rodgers, 2016, 2019; Rodgers et al., 2016; Sims et al., 2024)) which is based on a nationally-representative household probability sample. This dataset includes a broad range of psychological and health-related measures collected from 12,686 adolescents identified from 8,770 households on December 31, 1978. Respondents are surveyed biennially. Extensive information on the publicly-available data, sampling process, and measures is located on the Bureau of Labor Statistics (BLS) maintained website: https://nlsinfo.org/

To conduct this study using the requisite within-family information, we employed kinship pairs to support the genetically informed analysis. Our research team has completed a multi-year project to reliably and validly identify the NLSY kinship pairs (Rodgers et al., 2016) using both indirect and direct ascertainment of relatedness, resulting in a kinship classification (e.g., twins, full siblings, half-siblings, cousins, etc.) for approximately 95% (N=5,302 kinship pairs, 4006 of those full siblings) of the NLSY79 kinship pairs.

#### **Measures**

**Personality Measures.** In 2014, the NLSY79 subjects were administered the Ten-Item Personality Inventory (TIPI; Gosling et al., 2003). The TIPI is a self-report measure with two items per Big Five construct. Respondents indicated how well pairs of adjectives describe themselves (1 *Disagree strongly to* 7 *Agree strongly*). Although the TIPI is a brief-form measure with modest reliability, other NLS surveys have found it consistent with its slightly longer-form cousin, the mini-IPIP (Bureau of Labor Statistics, 2018; Donnellan et al., 2006). Descriptive statistics are presented in Table 1.

**SES Measures.** Two components of SES were used: highest grade completed and total net family income (TNFI). Highest grade completed was measured in years of education completed and ranged from 0 years to 20 years at age 50. All participants with more than 20 years of education (e.g., MD/PhDs) were truncated to a score of 20 in the released data. TNFI reflected the net household earnings. We converted all incomes into 2014 dollars using the Consumer Price Index (Crawford et al., 2016).

**Health Measures.** As respondents passed age 50, respondents were administered health “modules." Administration began in 2008 when the oldest respondents (i.e., those born in 1957 and 1958) reached 50. Administration of the health modules continued biennially until the youngest respondents reached age 50 in 2016. Approximately half of the subjects took the Health 50 at age 51.

***Short Form Health Survey.*** Th**e** 12-Item Short-Form Health Survey (SF-12; Ware et al., 1995), contains a norm-referenced physical component summary score (PCS). Higher scores correspond to greater health. Scores range from 0 to 100, with a population mean of 50 and a standard deviation of 10.

***Depression Scale****.* During this module, a 7-item version of the Center for Epidemiological Studies Depression Scale (CES-D; see Levine, 2013 for psychometric properties) was also administered. For each item, participants indicated how often they experienced the prompt during the past week (e.g., “I felt that everything I did was an effort.”). Responses ranged from 0 (*Rarely/None of the time)* to 3 (*Most/All of the time)*. Total scores were summed responses, ranging from 0 to 21.

### China Family Panel Survey (CFPS)

#### **Data**

The China Family Panel Survey (CFPS) is a comprehensive longitudinal study initiated in 2010 that provides insights into a variety of domains, including education, health, and social relationships (Xie & Hu, 2014). The survey, conducted biennially, is based on a nationally-representative sample from mainland China. As of 2018, it had surveyed 58,504 individuals from numerous households. The survey is publicly available and maintained by the Institute of Social Science Survey (ISSS) at Peking University. Detailed information about the CFPS, including its methodology and measures, is accessible on their website: https://www.isss.pku.edu.cn/cfps/en/

CFPS has an accurate and detailed record of kinship relationships both inside the family and throughout family changes. Following the strategy implemented by Rodgers and colleagues (2016), we developed preliminary kinship links using similar indirect and direct methods of determining relatedness. Based on the 2018 survey, we identified 116,909 unique relationships, from parent-child relationships to cousins in the CFPS-2018 dataset (Lyu & Garrison, 2022).

#### **Measures**

**Cognitive Ability**. Cognitive ability was assessed using a sequence of 24 progressively challenging mathematical problems, including addition, subtraction, multiplication, division, exponents, logarithms, trigonometric functions, sequences, permutations, and combinations (Institute of Social Science Survey, 2022). Question starting points varied based on the interviewee’s education level: for elementary education or below, questions began at number 1; for middle school level, at number 13; and for high school or higher, at number 19.

**Highest Grade**. We used the highest grade completed as a proxy measure of socioeconomic status (SES). Grades ranged from zero to 23 years.

**Depression**. Depression was measured using 9 items selected and translated from the Center for Epidemiologic Studies Depression Scale (CES-D; Radloff, 1977; Xu et al., 2021). Numerous studies have validated the CES-D's reliability and validity across diverse contexts (Zhang et al., 2012), cultures (Wang et al., 2013), and racial and ethnic groups (Kim et al., 2011).
