## Supplementary material for "Sibling Models Can Test Causal Claims without Experiments: Applications for Psychology": Tutorial

Supplemental Material

This document explains how to reproduce the analyses included in the manuscript “Sibling Models Can test Causal Claims without Experiments: Applications for Psychology”.

If you are familiar with version-control using git and GitHub, the easiest way to replicate our analysis is by cloning the project repository hosted at [github.com/R-Computing-Lab/target-causal-claims](https://github.com/R-Computing-Lab/target-causalclaims). Once cloned, you can open your favorite R IDE (e.g., RStudio or VSCode), and run the following:

*# Install renv if needed*
**if** (!require("renv")) install.packages("renv")
*# Retrieve the packages used in the analysis*
renv::restore()
*# Trigger the targets pipeline*
targets::tar_make()

This will recreate our analyses using the [targets](https://docs.ropensci.org/targets/) package, a pipeline toolkit for R by Will Landau (Landau 2021). Calling targets::tar_make() will trigger the pipeline, performing all data cleaning and modeling described in our paper. If you wish to replicate our analyses without cloning the repository from GitHub, please see the detailed walk-through for one of the studies below (the procedure is similar for all).

For this example, we are looking at whether neuroticism causally influences mental health. The first thing to do is load the packages we need for our analysis and define a utility function to reverse code survey items (necessary for combining measures from the TIPI).

library(tidyverse)
library(NlsyLinks)
library(discord)
library(gtsummary)

*#' Reverse Code Survey Items*
*#'*
*#' @description From https://github.com/svmiller/stevemisc/blob/master/R/revcode.R*
*#' @param x A vector of survey items*
reverse_code <- **function**(x) {
 len <- length(na.omit(unique(x)))+1
 return((x*-1) + len)
}

The next step is to read in and process the data. Please see the code comments for specifics on individual steps.

*# Set the url for neuroticism data*
neuroticism_data_url <- "https://raw.githubusercontent.com/R-Computing-Lab/target-causalclaims/main/data/neuroticism.csv"

*# Read in the raw data*
processed_neuroticism_data <- read_csv(
 neuroticism_data_url
) %>%
 *# Select the variables, renaming them according to the fields from NLSY*
 select(
 case_id = R0000100,
 sample_id = R0173600,
 race = R0214700,
 sex = R0214800,
 depression = H0013301, *# H0013301 is depression at age 50*
 anxious_upset = T4998603,
 calm_stable = T4998608
 ) %>%
 *# Filter the data frame so we only have values greater than zero in the columns 'depression_score', 'anxious_upset', and 'calm_stable'*
 filter(
 if_all(
 .cols = c(depression, anxious_upset, calm_stable),
 **function**(value) value >= 0)
 ) %>%
 *# Define neuroticism and scale relevant measures*
 mutate(
 depression = scale(depression)[,1],
 *# Reverse Coding calm/stable measurement from TIPI and adding with*
 *# anxious/upset*
 neuroticism = scale(
 reverse_code(calm_stable) + anxious_upset
 )[,1]
 )

In order to perform the discordant kinship regression, we need to properly define familial relationships for NLSY subjects. This is easily done using the the NlsyLinks R package(Beasley et al. 2016), the methods of which are described in (Rodgers et al. 2016).

single_entered_neuroticism <- processed_neuroticism_data %>%
 mutate(
 SubjectTag = case_id * 100
 ) %>%
 CreatePairLinksSingleEntered(
 *# Get familial links from the NLSY79 dataset for full siblings*
 linksPairDataset = Links79PairExpanded %>%
 filter(
 RelationshipPath == "Gen1Housemates" & RFull == 0.5
 ),
 linksNames = c(
 "ExtendedID",
 "R",
 "RelationshipPath"
 ),
 outcomeNames = c(
 "depression",
 "neuroticism",
 "sex",
 "race"
 ),
 subject1Qualifier = "_s1",
 subject2Qualifier = "_s2"
 ) %>%
 *# NAs in outcome variable cannot be present for discordant regression to work*
 drop_na(
 contains("depression")
 ) %>%
 as_tibble() %>%
 rename(
 subject_tag_s1 = SubjectTag_S1,
 subject_tag_s2 = SubjectTag_S2,
 extended_id = ExtendedID,
 relationship_path = RelationshipPath
 )

With this linked data, we can fit the discordant regression as follows:

neuroticism_fit <- discord_regression(
 data = single_entered_neuroticism,
 outcome = "depression",
 predictors = "neuroticism",
 sex = "sex",
 race = "race",
 pair_identifiers = c("_s1", "_s2")
 )

And view an aesthetically pleasing table of regression results:

neuroticism_fit %>%
 tbl_regression() %>%
 add_glance_source_note() %>%
 modify_header(
 statistic ~ "**t-statistic**",
 p.value ~ "**p-value**"
 )

| Characteristic | Beta | t-statistic | 95% CI^1^ | p-value |
| --- | --- | --- | --- | --- |
| depression_mean | 0.83 | 39.1 | 0.78, 0.87 | <0.001 |
| neuroticism_diff | 0.05 | 4.73 | 0.03, 0.07 | <0.001 |
| neuroticism_mean | -0.03 | -1.40 | -0.07, 0.01 | 0.2 |
| sex_1 | 0.03 | 1.05 | -0.03, 0.09 | 0.3 |
| race_1 | -0.08 | -3.99 | -0.12, -0.04 | <0.001 |
| sex_2 | -0.06 | -1.87 | -0.11, 0.00 | 0.061 |
| ^1^CI = Confidence Interval | | | | |
| R² = 0.466; Adjusted R² = 0.464; Sigma = 0.671; Statistic = 296; p-value = <0.001; df = 6; Log-likelihood = -2,082; AIC = 4,180; BIC = 4,225; Deviance = 917; Residual df = 2,039; No. Obs. = 2,046 | | | | |

Our GitHub repository contains both the [data](about:blank) for the other questions addressed in our paper and [commented code](https://github.com/R-Computing-Lab/target-causalclaims/blob/main/R/utils_process-data.R) for the specific variable transformations.
